# Estimates of the global burden of Congenital Rubella Syndrome, 1996-2019

**DOI:** 10.1101/2023.04.19.23288818

**Authors:** Emilia Vynnycky, Jennifer K Knapp, Timos Papadopoulos, Felicity T Cutts, Masahiko Hachiya, Shinsuke Miyano, Susan E Reef

## Abstract

**Background:** Many countries introduced rubella-containing vaccination (RCV) after 2011, following changes in recommended World Health Organization (WHO) vaccination strategies and external support. The full impact of these introductions is unknown as previous estimates of the global burden of Congenital Rubella Syndrome (CRS) considered the period 1996-2010.

**Methods:** We updated a previously-published literature review to identify rubella seroprevalence studies among unvaccinated populations. These were used in an age-structured transmission model, including routine and campaign vaccination coverage to estimate the CRS incidence during 1996-2019 in each country, each region and globally.

**Findings:** For 2019, the highest CRS incidence was estimated for the WHO African (AFR) and Eastern Mediterranean (EMR) regions (64 (95% CI: 24-123) and 27 (95% CI: 4-67) per 100,000 live births respectively), where nearly half of births occur in countries that have introduced RCV.. In regions elsewhere, where over 95% of births occurred in countries which had introduced RCV, the estimated CRS incidence was low (<1 (95% CI: <1-8) and <1 (95% CI: <1-12) per 100,000 live births in the South East Asian (SEAR) and Western Pacific (WPR) regions respectively, and similarly in Europe and the Americas). The estimated number of CRS births globally declined by approximately two thirds from 100,000 (95% CI: 54,000-166,000) in 2010 to 32,000 (95% CI: 13,000-60,000) by 2019, with the biggest falls in SEAR and WPR.

**Interpretation:** The introduction of RCV in SEAR and WPR led to dramatic regional and global reductions in the CRS incidence since 2010. Introducing RCV in the remaining countries and maintaining high RCV coverage can result in further reductions.

**Funding:** Gavi the Vaccine Alliance via the Vaccine Impact Modelling Consortium (VIMC). VIMC is jointly funded by Gavi the Vaccine Alliance and the Bill and Melinda Gates Foundation (BMGF grant number: OPP1157270).

## Introduction

Congenital Rubella Syndrome (CRS) is associated with significant disability, including congenital heart defects, cataracts, hearing impairment and developmental delay(1). When a woman is infected with rubella in early pregnancy, the infant may be born with CRS. The CRS burden is typically low in countries where coverage with rubella-containing vaccines (RCV) is high. Previous estimates(2) suggest that during 1996-2010, the number of children born globally with Congenital Rubella Syndrome (CRS) decreased modestly from 119,000 to just over 100,000. Much of the estimated CRS burden in 2010 was in Africa and South East Asia where rubella vaccination had not yet been introduced in 96% and 64% of countries(2). Since 2010, the World Health Organization (WHO) recommended all countries introduce rubella vaccine and suggested a preferred strategy of a campaign targeting a wide-age range(3), followed immediately by inclusion in the childhood immunization schedule. Gavi has also begun funding the introduction of RCV in many low income countries(4). Forty two countries have since introduced RCV and new estimates of the burden of CRS are needed to quantify the impact of their introduction(2).

Updated estimates of the pre-COVID era CRS burden are also needed for monitoring the impact of pandemic-related disruption. Recent estimates suggest that the COVID pandemic has disrupted immunization delivery services, with approximately 27.2 (95% CI: 23.4-32.5) million children missing their first dose of measles-containing vaccine (MCV) in 2019(5) and an estimated 7.9% reduction in the global MCV coverage in 2020 compared to that expected. Since RCV is administered together with MCV, prolonged disruption of immunization services could lead to increasing proportions of girls still being susceptible to infection when reaching child-bearing age, and hence an increased CRS burden.

In this paper, we update a previously-published literature review of rubella seroprevalence studies(2) and estimate the global CRS burden during 1996-2019 using mathematical modelling.

## Methods

### Data sources

#### Literature search to identify seroprevalence data

Our previous systematic review(2) covered 1990-December 2011. In our updated review, we identified age-stratified rubella seroprevalence data published between January 2012 and September 2020. We scanned citations in published papers and published(6) and unpublished literature reviews (Winter et al, personal communication) and a co-author’s (JK Knapp) archive for additional datasets. We also used an unpublished dataset provided by CDC from a representative population in Indonesia (S Reef, personal communication).

We followed previously-published methods (2, 7) to identify datasets from the literature search, reviewing the abstracts to identify potentially relevant articles, before reading the publication in full and extracting age-specific numbers of seropositive and seronegative individuals from eligible datasets, by sex where possible. “Child-bearing” age was assumed to be 15–44 years and equivocal rubella antibody results were interpreted as seropositive. Datasets were eligible for inclusion if they were considered unbiased(2) and collected before RCV had been introduced. For countries without data pre-dating the introduction of RCV, subsequent data collected in age groups that had remained unvaccinated were also eligible. Details of the search and eligibility criteria are provided elsewhere(2).

#### Demographic data

The total population size, age-specific numbers of females for 1996-2019 by single year and age, and age-specific fertility rates were extracted for each country from UN population databases(8). Fertility rates were available for five year age groups and time periods (1995-2000, 2000-2005, 2005-2010, 2010-2015 and 2015-2020). Annual values for each five year age group were interpolated from the corresponding period. For each country, the total number of live births for 1996-2019, the crude birth rate and age and sex-specific survival data for the period 2015-2020 were also extracted from UN population databases(8).

#### Vaccination data

We used UN population estimates of the annual number of live births in each country and WHO region to calculate the annual percentage of all live births in each WHO region during 1996-2019 which occurred in countries which had introduced RCV.

Countries have submitted annual vaccination coverage data to WHO since 1980(9). Since 2000, WHO and the United Nations Children’s Fund (UNICEF) jointly review these and available special survey data to obtain the WHO-UNICEF coverage estimates (WUENIC)(9). For 53 countries reporting having RCV in the national immunization schedule, but lacking RCV coverage data for the relevant years, we assumed that RCV coverage equalled the WUENIC estimate(10) for the first and second doses of MCV (MCV1 and MCV2) or the reported coverage if this was unavailable. Alternative coverage sources were identified if neither was available. For remaining gaps, the coverage data were interpolated (Supplement Table A).

Historical data on the target population and the estimated coverage for periodic mass RCV supplementary immunization activities (“SIAs”) available from WHO(11) and elsewhere (Supplement, Section A) were also used. For countries known to vaccinate adolescent girls for given years, we used published coverage data where possible (Supplement, Section A); otherwise, we assumed 50% coverage. In previous analyses(2), assuming either 10% or 90% coverage for adolescent girls did not substantially affect global burden estimates from 1996.

Reliable estimates of post-partum or private (but not public) sector vaccination were unavailable. Missing SIAs were supplemented from publications (Supplement, Section A). One country (Uganda) reportedly introduced RCV via an SIA in October 2019. Since this would not have greatly affected the burden in 2019, for simplicity, for Uganda, we assumed zero RCV coverage for 2019.

### Mathematical modelling

#### Overview

We first used catalytic models(12) to analyse the seroprevalence data identified in the literature review. The resulting pre-vaccination force of infection (rate at which susceptibles are infected) estimates were used to calculate age-dependent contact parameters, which were then included in an age-structured dynamic transmission model.

#### Catalytic modelling

The seroprevalence datasets identified in the updated literature review were analysed using four catalytic models (labelled A, B, C, D), as described previously(2, 13) to calculate the average age-specific force of infection, before RCV had been introduced (Supplement, section B). The catalytic models assumed that the pre-vaccination force of infection differed between those aged ≤13 and >13 years, and estimates from the most plausible model were used in CRS burden calculations(2, 13). For one setting (Laos) lacking data pre-dating the introduction of RCV, we used data collected several years after an SIA(14), and restricted the catalytic modelling to data from unvaccinated age groups, and assumed a 50% reduction in the force of infection after the SIA. Alternative assumptions for this reduction did not affect the pre-vaccination force of infection estimated among adults(14).

As in previous analyses, confidence intervals (CI) on the force of infection and (where applicable) the sensitivity of the assay for each dataset and catalytic model were generated using 1000 bootstrap-derived-seroprevalence datasets(2) using non-parametric bootstrap for binary data, based on 1000 bootstrap datasets, following Shkedy et al(15)

#### The transmission model

The transmission model is similar to one used previously (2, 13). The population is stratified into those with maternal immunity (lasting 6 months), susceptible, pre-infectious, infectious and immune through either vaccination or natural infection. The population was further stratified by sex into single year age groups, following the approach of Schenzle(16). In contrast with the model used previously, the population was aged 0-99 years and the same general model was used for all countries, irrespective of whether they had introduced vaccination. We used country-specific birth rates and age- and sex-specific death rates fixed at 2015-2020 levels, with the latter calculated using UN population survival data(8). The model’s differential equations and model diagram are in the supplement to reference (2).

Following previous methods, the force of infection in the transmission model changes over time and is calculated using the number of infectious individuals and the effective contact rate. Contact is described using the following matrix of “Who Acquires Infection From Whom”, whose general structure is based on contact survey data(17):

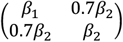

The effective contact rate differs between <13 and ≥13 year olds. For a given country, *β*_1_ and *β*_2_ are calculated from the pre-vaccination force of infection in <13 and ≥13 year olds, as estimated by fitting catalytic models to country-specific age-stratified rubella seroprevalence data(2) (see above).

For each country, the transmission model was run using 1000 values for the pre-vaccination force of infection, vaccine efficacy, vaccine coverage and risk that a child born to a mother who was infected with rubella whilst pregnant had CRS, which were varied in the same range as that used previously(13) (Supplement, Table B). The method used to compile the 1000 values for the pre-vaccination force of infection for a given country depended on the number of seroprevalence datasets available for that country (Table 1).

**Table 1:**
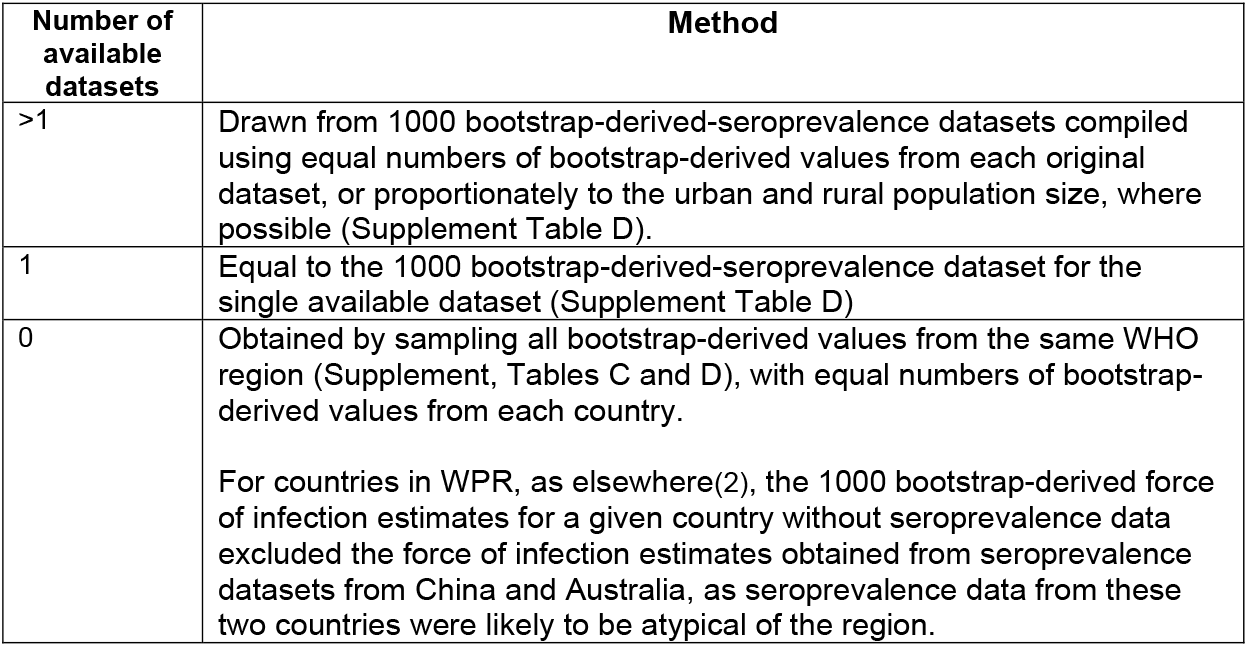
Method for compiling 1000 values for the pre-vaccination force of infection for a given country according to the number of seroprevalence datasets available for that country. Further details of the actual datasets used are in Supplement Table C

The transmission model was used to calculate the CRS incidence per 100,000 live births and number of CRS cases born annually during 1996-2019, for each country, WHO region, and globally (Supplement, Section D). We also computed the number of CRS cases in AFR, EMR, SEAR and WPRO and globally if RCV had not been introduced in these areas since 2010. We subtracted the estimated number of CRS cases obtained using the implemented RCV coverage from these numbers to obtain the number of CRS cases that were averted through introducing RCV after 2010. The 95% range of all model outputs, approximating the 95% confidence intervals, were calculated as the 95% range of the 1000 model runs. As previously, given China’s large population size, the regional incidence for WPR was calculated with and without excluding China.

### Role of the funding agency

The funding agency had no role in either the study design, collection, analysis or interpretation of the data, writing of the manuscript or the decision to submit the paper.

## Results

### Literature search and analyses of seroprevalence data

Figure 1 summarizes the literature search results. After de-duplication and excluding ineligible studies, we identified 21 potential serological datasets for estimating the average pre-vaccination force of infection. Five additional datasets were identified from scanning reference lists or conducting searches related to the identified studies, besides twelve datasets predating 1990 from a co-author’s (JKK) archive. Conferring with an unpublished literature review (Winter *et al, personal communication*) led to no further datasets being identified. After fitting catalytic models, the selected model for one dataset (Nigeria (Keffi(18)) had implausibly low force of infection estimates (Supplement, Table E) and was dropped from our analyses.

**Figure 1:**
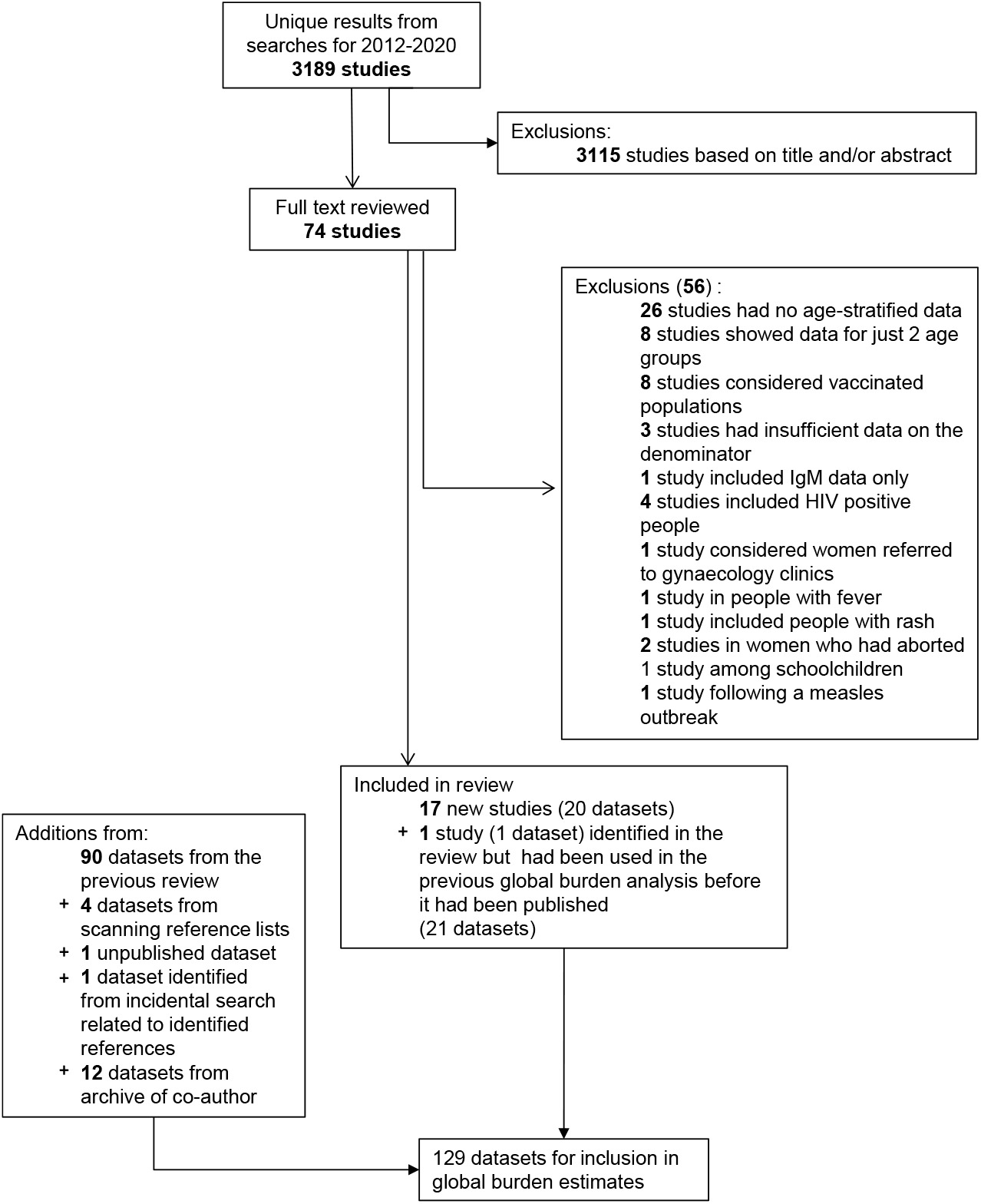
Results of the literature search for age-specific serological data for estimating the force of infection before the introduction of rubella vaccination. The small discrepancy between the number of datasets reported here for the previous review is due to the latter counting several datasets stratified by urban and rural locations as one dataset. In this figure, we refer to a publication as a study.

After including the datasets from the previous review(2) and one unpublished study from Indonesia provided by CDC (Susan Reef, *personal communication*), 129 datasets from 70 countries were available for estimating the global CRS burden (Table 2).

**Table 2:** Summary of the serological datasets collected which were used to estimate the global burden of CRS. Studies not used in the previous update are in bold font. The references can be found in the Supplement.

|  | **Africa** | **Americas** | **E Mediterranean** | **Europe** | **SE Asia** | **W Pacific** |
| --- | --- | --- | --- | --- | --- | --- |
| **No. countries represented** | 19 | 12 | 12 | 12 | 5 | 10 |
| **No. of countries in region** | 47 | 39 | 22 | 51 | 11 | 27 |
| **Datasets** | **Algeria, 2005-7**; Benin, 1993; **Burkina Faso, 2007-8**; **Cameroon (Bafoussam), 2016**, **Cameroon (Yaounde), <2018**; Congo, <1991; Cote d'Ivoire, 1975 & 1985-6; **Democratic Republic of the Congo (Kikwit, Mikalayi, Tshikapa, Vanga), 2008-9**; Ethiopia, 1981 & 1994; **Ethiopia (Amhara), 2015-17**; **Ethiopia (Hawassa), 2016**; Gabon, 1985; Ghana, 1997; Kenya, 1996-9 (Kilifi); **Kenya (Eldoret), 2005**; Madagascar, 1990-1995; Mozambique, 2002; **Namibia, 2010**; Nigeria, <1978, <2002 & 2007-8; **Nigeria (Kaduna), 2011-12**; **Nigeria (Ilorin ), 2012**, **Nigeria (Maiduguri), 2013**; **Nigeria (Kaduna), 2015**; Senegal, 1996-2001; South Africa, 2003, **South Africa (Soweto), 2014-16**; **Tanzania (Mwanza), 2012-13**; Zambia, 1979-80 | Argentina, 1967-8 (urban & rural) & 1981 (Mar de Plata); Brazil, 1967-8, 1987 & 1996-8; Canada, <1967; Chile 1967-8 (Santiago & rural); **Chile (Santiago), 1983**; Haiti, 2003; Jamaica, 1967-8 (Kingston & rural); Mexico, 1987-88 & 1989; Panama 1967-8 (Panama City & rural); Peru, 1967-8 (Lima & rural) & 2003; Trinidad 1966-7, 1967-8 (Port au Spain & rural); Uruguay, 1967-7 (urban and rural); USA <1967 (Atlanta & Houston) | Bahrain, 1981; **Egypt (Cairo), 1973**; Iran, 1993-95; Jordan, 1982-3; Kuwait, <1978; Lebanon, 1980-1; Morocco, 1969-70; Pakistan, <1997, 1999-2004; Saudi Arabia, 1989 & 1992-93; **Sudan (Khartoum), 2015-16**; **Sudan (Khartoum), 2016**; Tunisia, <1970; Yemen, 1985 & 2002-03 | Czech Republic, <1967 & **Czech Republic, 1984 (Prague)**; Denmark, <1967 & 1983; East Germany, 1990; England, <1967 & 1986-7; Finland, 1979; France, <1967; **Kyrgyzstan, 1968-70** and 2001; **Poland, 1969, 1973, 1979 (urban), 1979 (rural), 1982 (urban), 1982 (rural)**; Romania, <1989; **Spain, 1969-71**; **Switzerland, 1985**; Turkey, 1998, 2003-04 & 2005. | Bangladesh, 2004-5; India, 1968 (urban & rural Delhi), 1972-3 (Chandigarh & Lucknow), 1976 (Calcutta), <1987 (Delhi), <1990 (Delhi), 1999-2000 (urban and rural Vellore); **India (Kerala), 2016**; **India, 2017**; **Indonesia, 2007 (S Reef, personal communication)**; Nepal, 2008, Thailand, 1978 | Australia (<1967?); **Cambodia, 2012**; China (1979-80); Fiji, <1973; Japan, <1967 (Sapporo & Ohtsu); **Laos, 2014**; Malaysia, <1972; Singapore, 1975-79, Taiwan, 1984 & 1984-6; Central Vietnam, 2009-2010 |
| **# datasets (# with SS+ >1000)** | 35 (8) | 27 (4) | 16 (7) | 24 (8) | 15 (5) | 12 (7) |

\* The year in which the study was carried out is not known for several studies. For these studies, the table includes "<" followed by the year of publication. \* SS=sample size

The percentage of countries with data varied between regions, coming from up to 55% of the constituent countries in a region. The dataset size and quality also varied. WPR had the largest percentage of datasets which had a sample size exceeding 1000 individuals (58%, 7 out of 12 datasets), as compared with 23% (8/35) for AFR and 15% (4/27) for AMR.

### CRS incidence

For 2019, the highest regional CRS incidence (64 (95% CI: 24-123) per 100,000 live births) was estimated for AFR, followed by EMR (27 (95% CI: 4-67) per 100,000 live births) (Figure 2). The estimated CRS incidence was very low elsewhere, for example, <1 (95% CI: <1-8) and <1 (95% CI: <1-12) per 100,000 live births in SEAR and WPR respectively. Almost all the livebirths in the regions with a low CRS incidence occurred in countries which had introduced RCV (Figure 2).

**Figure 2:**
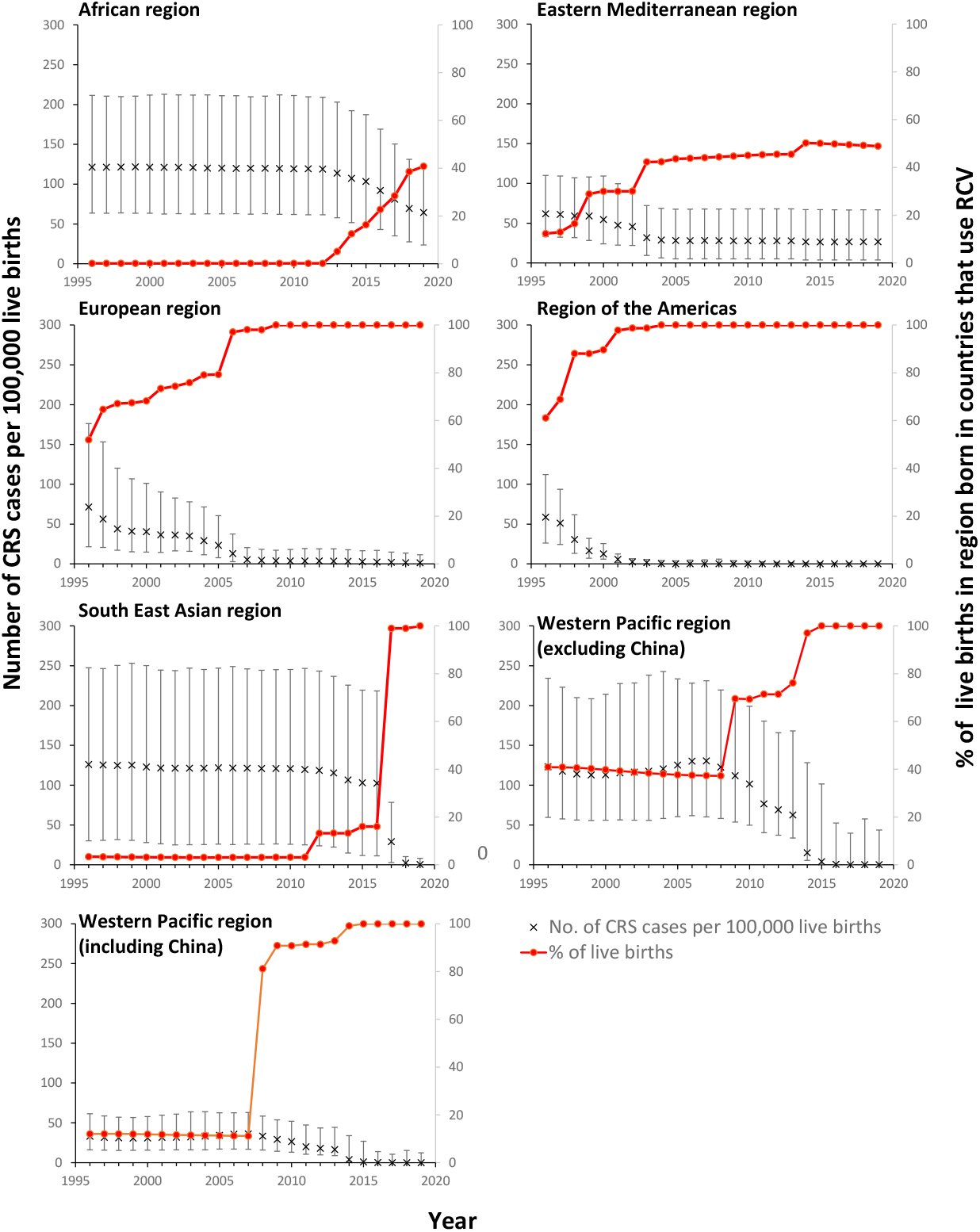
Estimates of the number of CRS cases per 100,000 live births since 1996 for each WHO region and the percentage of live births in each region occurring in countries which had introduced RCV. The error bars show the 95% range (CI).

For AFR, SEAR and WPR, large declines in the CRS incidence were estimated after 2010 (Figure 2). These declines corresponded to increases in the percentage of the regional live births occurring in countries which had introduced RCV. For AFR, the CRS incidence was stable until 2012, before decreasing from 119 (95% CI: 62-209) to 64 (95% CI: 24-123) per 100,000 live births by 2019. The incidence dropped dramatically in SEAR from 102 (95% CI: 11-218) to <1 (95% CI: <1-8) per 100,000 live births during 2016-2019, having been stable before 2016, with similar reductions in WPR, where it decreased slightly from 2008 and more dramatically from 2013. There was little change in EMR since 2003. By 2019, 40% and 50% of the estimated live births in AFR and EMR respectively were in countries which had introduced RCV, as compared with 100% in other regions.

The 22 countries that had not yet introduced RCV by mid 2019(19) were in AFR (17, 17% of global live births) and EMR (5, 7% of global live births), representing 60% and 50% of the livebirths in each respective region. Figure 3 shows the estimated CRS incidence in 2019 in these countries. For the seventeen countries in AFR, the estimated CRS incidence was around 100 per 100,000 live births, ranging from 36 (95% CI: 2-93) per 100,000 live births in South Africa to 130 (95% CI: <1-337) per 100,000 livebirths in Nigeria. For the five EMR countries, it ranged from 50 (95% CI: <1-173) to 91 (95% CI: <1-202) per 100,000 livebirths in Pakistan and Sudan respectively. The CRS incidence in the remaining countries which had introduced RCV remained low (<50 per 100,000 live births) (Supplement Figure S1).

**Figure 3:**
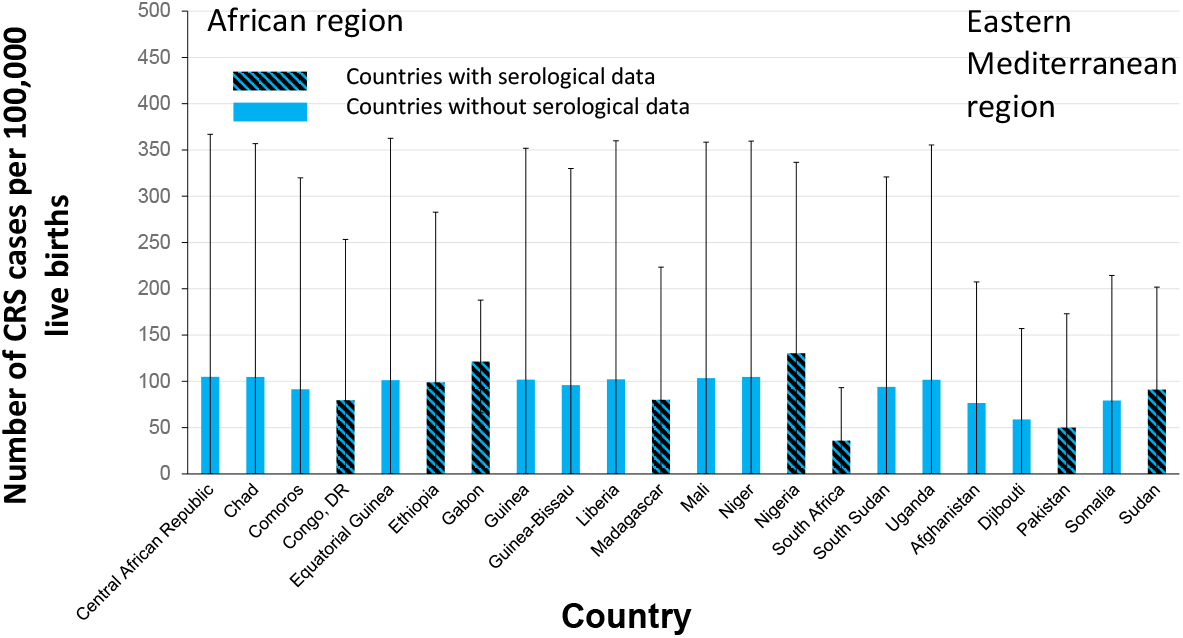
Estimates of the number of CRS cases per 100,000 live births in 2019 for countries which had not yet introduced RCV by mid-2019. The error bars show the 95% range (CI). The patterned and unpatterned bars reflect estimates for countries which had and did not have seroprevalence data respectively.

### Numbers of CRS cases

The estimated global annual number of CRS cases declined gradually during 1996-2016 from 121,000 (95% CI: 70,000-191,000) in 1996, reaching 100,000 (95% CI: 54,000-166,000) and 76,000 (95% CI: 33,000-134,000) by 2010 and 2016 respectively (Figure 4 and Supplement Table F). The estimated number fell more sharply since then, reaching 32,000 (95% CI: 13,000-60,000) by 2019.

**Figure 4:**
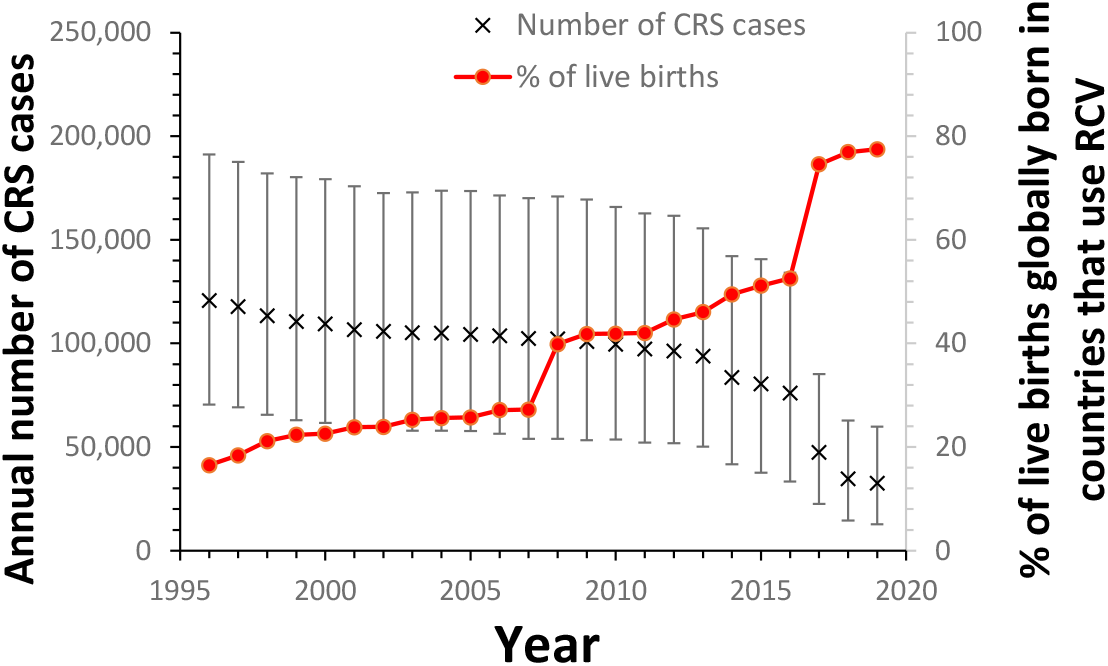
Estimates of the global number of CRS cases born annually during 1996-2019 and the percentage of the live births occurring globally in countries which had introduced RCV. The error bars show the 95% range (CI).

By 2019, the highest numbers of CRS cases were estimated for AFR (25,000 (95% CI: 9200-49,000), followed by EMR (5700 (95% CI: 870-14,000)) and SEAR (51 (95% CI: <1-1660)). The numbers elsewhere were <1 (95% CI: 0-90), 100 (95% CI: <1, 960) and <1 (95% CI: 0-3770) (AMR, EUR and WPR respectively).

Trends in annual numbers of CRS cases until 2019 differed between regions (Figure 5), reflecting differences in the population growth rate and timing of RCV introduction. In AFR, secular increases in the number of births, as noted previously(2), were estimated to lead to an increase in the number of CRS cases from 1996, from 29,000 CRS cases (95% CI: 15,000-53,000) to reach 39,000 (95% CI: 20,000-70,000) by 2012, before returning to a similar level to that estimated for 1996 by 2019. In EMR, the estimated annual number of CRS cases changed little after 2004, whilst in SEAR, it remained relatively unchanged until 2016, and subsequently declined rapidly.

**Figure 5:**
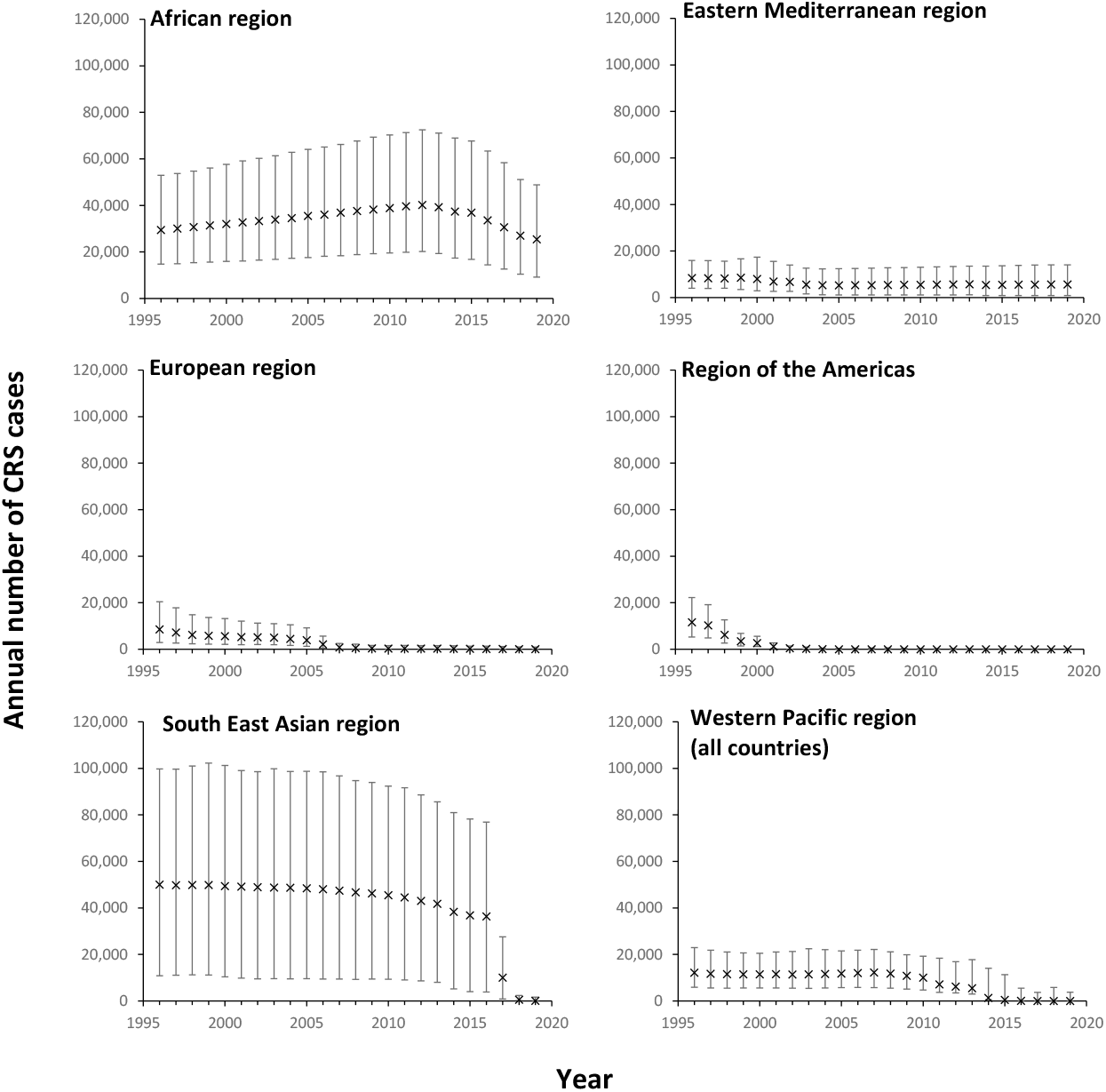
Estimates of the number of CRS cases born annually in each WHO region during 1996-2019. The error bars show the 95% range (CI).

By 2019, the highest average number of CRS cases (>1000 annually) were estimated in 7 countries: Democratic Republic of the Congo, Ethiopia, Niger, Nigeria, Uganda, Pakistan, Sudan (Figure 6). For eleven countries in AFR (Benin, Central African Republic, Chad, Guinea, Liberia, Madagascar, Mali, South Africa, South Sudan) and two countries in EMR (Afghanistan, Somalia), the estimated average annual number of CRS cases was in the range 100-999. For several countries, these estimates had very wide 95% CI, for example, <1 (95% CI: <1-3,600) for the Philippines.

**Figure 6:**
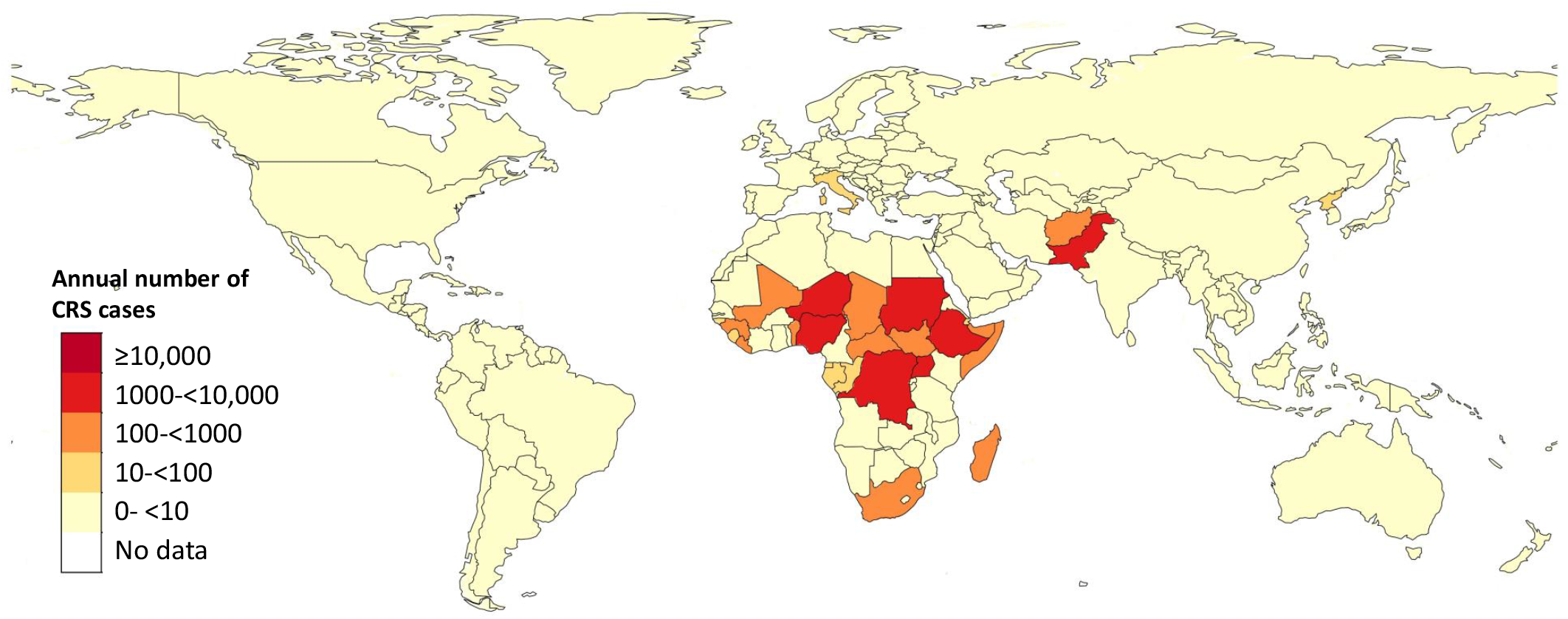
Estimates of the average number of CRS cases born annually in each country in 2019.

### Numbers of CRS cases averted by the introduction of RCV since 2010

Without the introduction of RCV since 2010, we estimate that the global CRS burden would have reached 96,000 (95% CI: 54,000-157,000) by 2019 (Table 3 and Supplement Figure S2). The corresponding numbers in AFR and SEAR were similar (44,000 (95% CI: 22,000-79,000) and 40,000 (95% CI: 8700-81,000) respectively); those in EMR and WPR were lower (6,000 (95% CI: 1200-14,000) and 5800 (95% CI: 3400-11,000) respectively) (Table 3 and Supplement Figure S2). Introducing RCV after 2010 averted an estimated 229,000 (95% CI: 131,000-368,000) CRS cases globally during 2011-2019, with the greatest number averted in SEAR (125,000 95% CI: 37,000-235,000) followed by 65,000 (95% CI: 37,000-111,000), 36,000 (95% CI: 21,000-59,000) and 1500 (95% CI: 400-3300) in AFR, WPR and EMR respectively (Table 3).

**Table 3:** Comparison between the estimated number of CRS cases in 2010, 2019 and during 2011-2019 with vaccination as implemented, and the numbers that might have been expected if vaccination had not been introduced after 2010. The final column shows the estimated number of CRS cases averted during 2011-2019 by the introduction of RCV after 2010.

|  | Year | Number of CRS cases |  | Total number of CRS cases during 2011-2019 |  | Number averted during 2011-2019 by RCV introduced after 2010 |
| --- | --- | --- | --- | --- | --- | --- |
|  |  | RCV as implemented | RCV not introduced after 2010 | RCV as implemented | RCV not introduced after 2010 |  |
| **African region** | **2010** | 38873<br>(19574,70294) | 38873 (19574,70294) | - | - | - |
|  | **2019** | 25454<br>(9193,48881) | 44146 (22018,79076) | 349510<br>(162083,648091) | 414344<br>(209074,746863) | 65370 (37446,111474) |
| **Eastern Mediterranean region** | **2010** | 5514<br>(1116,13043) | 5514 (1116,13043) | - | - | - |
|  | **2019** | 5660<br>(873,14073) | 5976 (1195,14348) | 55976<br>(9773,135945) | 57895<br>(11580,137674) | 1504 (417,3323) |
| **South East Asian region** | **2010** | 45444<br>(9316,92325) | 45444 (9316,92325) | - | - | - |
|  | **2019** | 51 (0,1662) | 40036 (8699,81023) | 297575<br>(50447,627451) | 422457<br>(89519,866180) | 124896 (37346,234769) |
| **Western Pacific Region** | **2010** | 10023<br>(4704,19261) | 10023 (4704,19261) | - | - | - |
|  | **2019** | <1 (0,3768) | 5829 (3412,11251) | 31362<br>(16089,114954) | 67706<br>(37839,152574) | 35741 (21030,59327) |
| **Global** | **2010** | 99665<br>(53554,165778) | 99665 (53554,165778) | - | - | - |
|  | **2019** | 32460<br>(12794,59818) | 96271 (53937,157435) | 743197<br>(392673,1254921) | 969653<br>(531016,1626758) | 228969<br>(130929,367995) |

## Discussion

We estimate that the number of CRS cases globally since 2010 has decreased by about two thirds, from 100,000 (95% CI: 54,000-166,000) to reach 32,000 (95% CI: 13,000-60,000) by 2019, with wide and overlapping confidence intervals. This reduction is largely due to rapid increases in population immunity after introducing RCV, resulting from the WHO preferred strategy to introduce RCV with a SIA covering a wide age range(3). We estimated dramatic (>90%) reductions in less than a five year period in SEAR and WPR, where, by 2019, 100% of births occurred in countries with RCV. Modest reductions were estimated for AFR and EMR, where 40 and 50% respectively of all births were estimated to occur in countries which had introduced RCV by 2019. RCV introduction after 2010 averted an estimated 229,000 (95% CI: 131,000-368,000) CRS cases globally during 2011-2019.

Most of the estimated burden in 2010 was in AFR and SEAR. We estimate that the burden in SEAR has reduced dramatically with the introduction of RCV including widespread SIAs into several populous countries, including Indonesia in 2017 and India by 2018. Similarly, the CRS burden declined in WPR in 2014 after RCV was introduced in Cambodia and Vietnam.

In contrast, the burden in AFR is estimated to have decreased only gradually, after RCV was introduced in less populous countries, such as Senegal (2012), followed by Ghana and Rwanda (2013), Burkina Faso and Tanzania (2014), Cameroon (2015), Eswatini, Gambia, Kenya, Namibia and Zambia (2016). The steady decline could continue, with RCV being introduced in Uganda late in 2019, which was not accounted for in these analyses.

Currently, the RCV introductions into the remaining – including the most populous - countries in AFR remains unscheduled. The CRS burden in EMR may change rapidly in future, since Pakistan introduced RCV through one of the world’s largest SIAs in late 2021(20) and other countries are finalising plans.

Our transmission model estimated rapid (>50%) reductions in CRS incidence soon after rubella vaccination is introduced. Such rapid reductions are consistent with observed data and with findings from other models investigating the introduction of RCV with SIAs (21). For example, a recent study(22) in five African countries, in which RCV was introduced through an SIA targeting children aged 9 months - 14 years in 2013 found a typical 48-96% reduction in the mean incidence of confirmed rubella cases after the SIA, compared to that beforehand.

Our estimates are subject to several limitations, as detailed elsewhere(2), including factors relating to the representativeness of the population surveyed in the seroprevalence data. For simplicity, we have not included the effect of correlations between vaccine doses and between vaccination received routinely and those received in a SIA. The correlation between the latter may be population-specific. For example, in one recent study, the percentage of children who had not received measles vaccination who subsequently received vaccination during an SIA was inversely affected by wealth status(23).

We may have overestimated the CRS burden in some settings by only including vaccination occurring through the routine schedule or during SIAs, given recent WHO recommendations(24) to give one dose of RCV to all unvaccinated or seronegative non-pregnant women, who are potentially identified at pre-marital screening, post-partum or when they contact the health system for other reasons. Conversely, we may have underestimated the impact of vaccinating children on transmission among adults by basing assumptions on the assortativity of mixing patterns on studies from Western settings, where adults may mix more assortatively than do adults in low and middle-income settings(25).

Our results rely on estimates of the pre-vaccination force of infection based on seroprevalence data from populations in which routine RCV vaccination had not been introduced. With the introduction of RCV in many countries, new studies collecting such data are increasingly rare. For example, our systematic review, which considered the same databases and search criteria as those used previously, identified fewer than 20 seroprevalence studies over the period 2012-2020 that were eligible for inclusion. However, as illustrated in previous analyses(14), a country with recent RCV introduction can collect seroprevalence data in unvaccinated age groups, from which we can estimate the pre-vaccination force of infection, after we make assumptions about the reduction in the force of infection resulting from vaccination.

Future trends in the CRS burden depend on many factors, including importation and variations in coverage. For simplicity the effects of importation of infection have not been included here and, as for measles and other infections(26), such importations could potentially lead to outbreaks, with the extent of sustained transmission depending on the population’s immunity levels. Variations in coverage, resulting from the ongoing COVID pandemic may have also affected the CRS burden. WUENIC data indicate that the global coverage of RCV decreased from 69% to 66%(27). Disruptions and delays to routine vaccination and the implementation of SIAs could cause increases in the proportion of women reaching child-bearing age still susceptible, which, coupled with ongoing transmission, could potentially lead to eventual increases in the CRS burden.

In conclusion, between 2010 and 2019, we estimate a reduction of over two thirds in the global CRS burden, with 229,000 (95% CI: 131,000-368,000) cases averted during 2011-2019, following the updated WHO introduction strategy, with a catch-up SIA and resources available through GAVI-funded initiatives. Our model estimated particularly dramatic (>90%) reductions in CRS incidence in SEAR and WPR in less than 5 years. With sustained coverage and the introduction of RCV into the remaining countries where it has not yet been introduced, further reductions are possible, making the long-term goal of eradication feasible. These hard-won achievements must not be destabilised by the consequences of the COVID pandemic.

## Data Availability

All data used in the present study are available upon reasonable request to the authors

## Acknowledgments

We are grateful to Amy Winter for sharing the results of her literature review.

## Data sharing

The data used in the analysis will be made available on a Github site when the paper is published.

## Disclaimer

The findings and conclusions in this report are those of the authors and do not necessarily represent the views of the Centers for Disease Control and Prevention.

## Author contributions

EV carried out the updated literature review, wrote and ran the code for estimating the force of infection, the transmission model and compiling the results, interpreted the results, wrote the paper.

JK compiled the vaccination coverage data, contributed to the literature review and to the interpretation and writing of the paper.

FC conducted a previous literature on which the current literature review is based, reviewed the updated datasets, contributed to the interpretation and writing of the paper.

TP contributed to the interpretation, generated figures and writing the paper.

MH contributed data, and contributed to the interpretation and writing of the paper. SM contributed data, and contributed to the interpretation and writing of the paper. SR contributed data and to contributed to the interpretation and writing of the paper. All authors had access to the data.

