## Supplementary material for "Estimates of the global burden of Congenital Rubella Syndrome, 1996-2019"

#### Table of contents

**A. Vaccination coverage data**……………………………………………………….………3

Table A: Countries and time periods for which RCV coverage for a given dose was estimated. One of four approaches was used: linear interpolation, constant gap, flat line and set value approaches.……………………………………………………………………………..4

**B. Analyses of additional seroprevalence datasets**……………………………………....5

**C. Parameter values in the transmission model**……………………………………….…..7

Table B: Summary of the basecase and ranges of the parameters used in the transmission model.........................................................................................................................7

Table C: Datasets used to set up 1000 force of infection bootstrap files for the WHO Regions. These bootstrap files were used to generate 1000 contact parameters for use in the transmission model to calculate the median and 95% range of the CRS burden for countries in a given region which did not have any pre-vaccination seroprevalence data (see main text). Note that these datasets had been accepted after performing the selection procedure described in section B. Table D provides further details of bootstrap data used for each country........................................................................................................................8

Table D: Summary of the bootstrap datasets used to establish the pre-vaccination force of infection for each country using the transmission model, using the WHO regional grouping to assign datasets for countries without serological datasets from before the introduction of RCV. See Table C for the datasets used to make up the bootstrap datasets........................8

**D: Equations for the CRS incidence and annual numbers of CRS cases**……...……..17

**E. Estimates of the pre-vaccination force of infection**………...……………………………19

Table E: Summary of the additional datasets that were identified since the previous systematic review, best-fitting values for the force of infection and (where appropriate) the sensitivity of the antibody assay for each catalytic model before the introduction of RCV. The values in parentheses reflect the 95% confidence intervals, obtained by bootstrapping. Several of the estimates were published in an interim update of the literature review(126) and are included here for completeness.…………………………………………………………………………………20

**F. Estimates of the CRS incidence**…………….………………………………………….33

Figure S1: Average estimates of the number of CRS cases per 100,000 live births in 2019 for all countries.…………………………………………………………..…………………33

Table F: The median CRS incidence per 100,000 live births and number of CRS cases born in each WHO region and worldwide in 1996, 2000, 2010 and 2019 and the percentage of the regional live births occurring in countries which had introduced RCV by these years. The numbers in parentheses reflect 95% confidence limits.........................................................................................................................34

Figure S2: Estimates of the annual number of CRS cases during 1996-2019 globablly, in the African, Eastern Mediterranean, South East Asian regions, as calculated using the RCV coverage as implemented and the number that might have been seen if there had been no new introductions of RCV after 2010. The shaded areas show the 95% confidence intervals.........................................................................................................35

#### A. Vaccination coverage data

The vaccination coverage data that were used in the model are described in the main text. Missing SIA or routine coverage data were further supplemented from publications(1-22). Data on coverage among adolescent girls came from publications where possible(23-25) (see main text for details).

Table A: Countries and time periods for which RCV coverage for a given dose was estimated. One of four approaches was used: linear interpolation, constant gap, flat line and set value approaches.

**Linear interpolation**

| ISO | Dose | Years |
|---|---|---|
| AND | 1st | 1989-1996 |
| AND | 2nd | 1991-1996 |
| ARG | 2nd | 1998-1999 |
| ATG | 2nd | 1997-2004 |
| AUT | 2nd | 1994-1999 |
| BHS | 2nd | 2001-2003 |
| BRA | 1st | 1992-1999 |
| CAN | 2nd | 2003,2005,2007,2008,2010 |
| DEU | 1st | 1988-1990 |
| GRC | 2nd | 1999-2006 |
| IRL | 2nd | 1995-1997 |
| ISL | F | 1982 |
| JPN | 2nd | 2006-2007 |
| KOR | 2nd | 1997-1999 |
| LBN | 2nd | 1996-1999 |
| MHL | 1st | 1989 |
| SGP | F | 1977-1980 |
| SVN | 2nd | 1999 |

**Fixed gap[%]**

| ISO | Dose | Years | Source year(s) of gap estimate |
|---|---|---|---|
| AND | 2nd | 1997-2006 | 2007 |
| ARE | 2nd | 1995-1999 | 2000 |
| BEL | 2nd | 1994-2005 | 2006 |
| BRN | 2nd | 1996-1999 | 2000 |
| BTN | 2nd | 2006-2007 | 2008 |
| CHE | 2nd | 1997-2004 | 2005 |
| CHL | 2nd | 1992-2003 | 2004-2018* |
| COL | 2nd | 1997-1999 | 2000 |
| CPV | 2nd | 2016 | 2017 |
| CRI | 2nd | 1992-2003 | 2008-2018* |
| CYP | 2nd | 1999-2006 | 2007-2018* |
| CZE | 2nd | 1995-1999 | 2000 |
| ESP | 2nd | 1995-2003 | 2004 |
| EST | 2nd | 1994-1999 | 2000 |
| FIN | 2nd | 1982-2015 | 2016-2018* |
| FRA | 2nd | 1996-2009 | 2010 |
| GBR | 2nd | 1996-1999 | 2000 |
| GRD | 2nd | 2000-2001 | 2002-2018* |
| HRV | 2nd | 1994-2000 | 2001 |
| ISL | 2nd | 1987-2002 | 2003-2010* |
| LUX | 2nd | 1995-2012 | 2013 |
| MEX | 2nd | 1998-1999 | 2000-2018* |
| MNE | 2nd | 1996-1999 | See Serbia dose 2 |
| NZL | 2nd | 1992-2010 | 2011 |
| PRT | 2nd | 1990-1999 | 2000 |
| QAT | 2nd | 1992-2001 | 2002 |
| RUS | 2nd | 1997-1999 | 2000 |
| SLV | 2nd | 2000-2002 | 2003 |
| SRB | 2nd | 1996-1999 | 2000 |
| SVK | 2nd | 1994-1999 | 2001 |
| SVN | 2nd | 1992-1998 | 2000 |
| SWE | 2nd | 1982-1999 | 2000-2018* |
| TON | 2nd | 2002 | 2003 |

**Flat line**

| ISO | Dose | Years | Source year(s) |
|---|---|---|---|
| AUS | F | 1989-1998 | 1988 |
| AUS | M | 1993-1998 | 1988 |
| BRN | F | 1988-1998 | 1987 |
| CYP | F [#] | 1972-1979 | 1980 |
| DEU | 2nd | 1986-1990 | 14%[$] |
| DEU | 2nd | 1992-1997 | 1998 |
| HKG | F | 1978-1979 | 1980 |
| IRL | F | 1980-1982 | 1983 |
| IRL | 2nd | 2000-2011 | Mid-point for 1999 & 2012 |
| IRL | 2nd | 2013-2018 | 2012 |
| KAZ | F | 2007-2009 | 2007 |
| LBY | 1st | 1993-2000 | 2001 (MCV2) |
| MCO | 2nd | 1996-2013 | 2014 |
| MYS | F | 2002-2009 | 2001 |
| MYS | 3rd [&] | 2016-2023 | 2015 |
| NZL | F | 1990-1991 | 1989 |
| NZL | F | 1979 | 1980 |
| PRT | F | 1987-1989 | 1986 |
| SGP | 2nd | 1982-1998 | 1981 |

**Set value**

| ISO | Dose | Years | Assumed value | Reasoning |
|---|---|---|---|---|
| CPV | 1st | 2010 | 25% | Introduced in Sept 2010 |
| JPN | F | 1977-1981, 1996 | 70% | Consistent with constant (67-77%) coverage during 1982-1994 |
| SGP | M | 1982-1990 | 100% | Mandatory conscription- Vx likely. |

[%] The coverage is calculated so that the absolute difference in the coverage between the first and second doses in the given year is fixed at the level observed in the years in column 7, unless otherwise stated. Consistent with steady coverage of the first dose.

[#] Adolescent

[$] Assumed to be at the level for the whole of Germany in 1990, multiplied by 0.797 (the factor by which the population in W Germany differed from the total population in Germany

* Calculated as the average annual difference between the first and second dose coverage for the period specified

[&] Provided for 7 year olds

#### B. Analyses of additional seroprevalence datasets

The methods used to estimate the force of infection for the seroprevalence datasets identified since the previous related analyses are described in (26). In brief, four catalytic models (A, B, C, D) were fitted to the age-stratified seroprevalence data to estimate the average annual "force of infection" among <13 and ≥13 year olds (i.e. the rate at which susceptible <13 and ≥13 year olds are infected), and the sensitivity of the antibody assay. The pre-vaccination force of infection was allowed to either differ for those aged <13 and ≥13 years (models A and B) or be identical for all ages (models C and D). The sensitivity of the rubella serological (antibody) assay was either estimated (models A and C) or assumed to be 100% (models B and D).

The criteria for selecting the force of infection for further use are described in (26). In brief, the force of infection estimates were selected in decreasing order of biological plausibility of the model, coming from model A, unless they met specific criteria (the force of infection was implausibly high (>600 per 1000 per year), zero in either age group, higher for older individuals than for children or its upper confidence limit was 100%). If this occurred, we used estimates from model B in preference to those from model C, and those from model C in preference to those from model D. If no model fitted the data convincingly, occurring when the best-fitting age-specific proportion seronegative passed through the 95% confidence limits of just one of the observed datapoints, the dataset was dropped from further analyses.

We included an additional criterion for countries for which the sensitivity of the assay was known to be high that model B was selected in preference to model A if all the other criteria were satisfied and the estimated sensitivity of the assay was 100%for model A, and the lower limit of the 95% confidence interval was implausibly low (less than 95%)(27). Table E: summarises the best-fitting values for the force of infection for the additional datasets for which the analyses have not yet been published.

#### C. Parameter values in the transmission model

Table B: Summary of the basecase and ranges of the parameters used in the transmission model.

| | **Base-case value** | **Values used in sensitivity analyses** | **Basis** |
|---|---|---|---|
| **Pre-vaccination force of infection (used to calculate contact parameters)** | Based on pre-vaccination seroprevalence data from the country (if available) or from the same WHO region otherwise. | 1000 bootstrap-derived values | See (26) and main text. |
| **Vaccine efficacy** | 95% | 85% to 99%, sampled from the truncated Beta distribution with parameters $\alpha$=33 and ß=2. | Plausible values |
| **Vaccination coverage** | See main text | 10% higher or lower each year than historical projections. | Plausible |
| **Risk of a child being born with CRS if the mother is infected during the first 16 weeks of pregnancy** | 65% | Sampled from the Gamma distribution with shape and scale parameters 37 and 56 respectively. | Lead to a median and 95% range of 65% and 47-88% respectively consistent with those from several studies(28-30) which, as found in a recent review(31) were likely to have been more reliable than those in other studies. |

Table C: Datasets used to set up 1000 force of infection bootstrap files for the WHO Regions. These bootstrap files were used to generate 1000 contact parameters for use in the transmission model to calculate the median and 95% range of the CRS burden for countries in a given region which did not have any pre-vaccination seroprevalence data (see main text). Note that these datasets had been accepted after performing the selection procedure described in section B. Table D provides further details of bootstrap data used for each country.

| **Region** | **Datasets** |
|---|---|
| African (AFR) | Algeria, 2005-7(32), Benin, 1993(33); Burkina Faso, 2007-8(34); Cameroon, 2016 (Bafoussam)(35) and <2018 (Yaounde) (36); Congo, <1991(37); Cote d'Ivoire, 1975(38) & 1985-6(39); Democratic Republic of the Congo, 2008-9 (Kikwit, Mikalayi, Tshikapa, Vanga) (27); Ethiopia, 1981(40) & 1994(41), 2015-17 (Amhara)[69], 2016 (Hawassa)(42) ; Gabon, 1985(43); Ghana, 1997(44); Kenya, 1996-9 (Kilifi)(45, 46), 2005 (Eldoret); Madagascar, 1990-1995(47); Mozambique, 2002(48); Namibia, 2010 (49); Nigeria, <1978(50), <2002(51) & 2007-8(52), 2011-12 (Kaduna) (53), 2012 (Ilorin) (54), 2013 (Maiduguri) (55), 2015 (Kaduna) (56); Senegal, 1996-2001(57); South Africa, 2003(58), 2014-16 (Soweto) (59); Tanzania, 2012-13 (Mwanza); Zambia, 1979-80(60), |
| American, excluding Caribbean (AMR, excl Caribbean) | Argentina, 1967-8 (urban & rural)(61), & 1981 (Mar de Plata)(62); Brazil, 1967-8(61), 1987(63) & 1996-8(64); Canada, <1967(65); Chile 1967-8 (Santiago & rural)(61) and 1983(66); Mexico, 1987-88(67) & 1989(68); Panama 1967-8 (Panama City & rural)(61); Peru, 1967-8 (Lima & rural)(61) & 2003(69); Uruguay, 1967-7 (urban and rural)(61); USA <1967 (Atlanta & Houston)(65). |
| Caribbean | Haiti, 2003(70), Jamaica, 1967-8 (Kingston & rural)(61), Trinidad 1966-7(71), 1967-8 (Port au Spain & rural)(61) |
| Eastern Mediterranean (EMR) | Bahrain, 1981(72); Egypt (Cairo), 1973(73); Iran, 1993-95(74); Jordan, 1982-3(75); Kuwait, <1978(76); Lebanon, 1980-1(77); Morocco, 1969-70(78); Pakistan, <1997(79), 1999-2004(80), 2Saudi Arabia, 1989(81) & 1992-93(82); Sudan, 2015-16 (Khartoum) (83), 2016 (Khartoum) (84); Tunisia, <1970(85); Yemen, 1985(86) & 2002-03(87) |
| European (EUR) | Czech Republic, <1967(65) & 1984 (Prague) (88); Denmark, <1967(65) &1983(89); East Germany, 1990(90); England, <1967(65) & 1986-7(91); Finland, 1979(92); France, <1967(65); Kyrgyzstan, 1968-70(93) & 2001(94); Poland, 1969(95), 1973(95), 1979 (urban)(96), 1979 (rural)(96), 1982 (urban)(95), 1982 (rural)(95); Romania, <1989(97); Spain, 1969-71(98); Switzerland, 1985(99); Turkey, 1998(100), 2003-04(101) & 2005(102). |
| South East Asian (SEAR) | Bangladesh, 2004-5(103); India, 1968 (urban & rural Delhi)(104), 1972-3 (Chandrigarh & Lucknow)(104), 1976 (Calcutta)(105), <1987 (Delhi)(106), <1990 (Delhi)(107), 1999-2000 (urban and rural Vellore)(108), 2016 (Kerala) (109), 2017 (110); Indonesia, 2007 (*S Reef, personal communication);* Nepal, 2008(111), Thailand, 1978(112) |
| Western Pacific (WPR), excluding China & Australia | Fiji, <1973(113); Japan, <1967 (Sapporo &Ohtsu)(65); Laos, 2014 (114); Malaysia, <1972(115); Singapore, 1975-79(116), Taiwan, 1984(117) & 1984-6(118); Central Vietnam, 2009-2010(119) |

Table D: Summary of the bootstrap datasets used to establish the pre-vaccination force of infection for each country using the transmission model, using the WHO regional grouping to assign datasets for countries without serological datasets from before the introduction of RCV. See Table C for the datasets used to make up the bootstrap datasets.

| Country | Bootstrap dataset used: |
|---|---|
| **Africa** | |
| Algeria | Algeria, 2005-7(32) |
| Angola | AFR |
| Benin | Benin, 1993(33) |
| Botswana | AFR |
| Burkina Faso | Burkina Faso, 2007-8(34) |
| Burundi | AFR |
| Cameroon | Cameroon, 2016 (Bafoussam) (35) & (Yaounde), <2018(36) |
| Cape Verde | AFR |
| Central African Republic | AFR |
| Chad | AFR |
| Comoros | AFR |
| Congo | AFR |
| Côte d'Ivoire | Cote d'Ivoire, 1975(38) & 1985-6(39) |
| Democratic Republic of the Congo | Democratic Republic of the Congo, 2008-9 (Kikwit, Mikalayi, Tshikapa, Vanga) (27) |
| Equatorial Guinea | AFR |
| Eritrea | AFR |
| Ethiopia | Ethiopia, 1981(40), 1994(41), 2015-17 (Amhara) (120), 2016 (Hawassa) (42) |
| Gabon | Gabon, 1985(43) |
| Gambia | AFR |
| Ghana | Ghana, 1997(44) |
| Guinea | AFR |
| Guinea-Bissau | AFR |
| Kenya | Kenya (Kilifi), 1996-9(45, 46), 2005 (Eldoret) (121) |

| | |
|---|---|
| Lesotho | AFR |
| Liberia | AFR |
| Madagascar | Madagascar, 1990-1995 (47) |
| Malawi | AFR |
| Mali | AFR |
| Mauritania | AFR |
| Mauritius | AFR |
| Mozambique | Mozambique, 2002(48) |
| Namibia | Namibia, 2010(49) |
| Niger | AFR |
| Nigeria | Nigeria, <1978(50), <2002(51) & 2007-8(52), 2011-12 (Kaduna) (53), 2012 (Ilorin) (54), 2013 (Maiduguri) (55), 2015 (Kaduna) (56) |
| Réunion | AFR |
| Rwanda | AFR |
| Sao Tome and Principe | AFR |
| Senegal | Senegal, 1996-2001 (57) |
| Seychelles | AFR |
| Sierra Leone | AFR |
| South Africa | South Africa, 2003 (58), 2014-16 (Soweto) (59) |
| South Sudan | AFR |
| Swaziland | AFR |
| Togo | AFR |
| Uganda | AFR |
| United Republic of Tanzania | Tanzania (Mwanza), 2012-13(122) |
| Western Sahara | AFR |
| Zambia | Zambia, 1979-80 (60) |
| Zimbabwe | AFR |

| **Americas** | |
|---|---|
| Antigua and Barbuda | Caribbean |
| Argentina | Argentina, 1967-8 (urban & rural)(61), & 1981 (Mar de Plata)(62) |
| Aruba | Caribbean |
| Bahamas | Caribbean |
| Barbados | Caribbean |
| Belize | Caribbean |
| Bolivia | AMR, excluding the Caribbean |
| Brazil | Brazil, 1967-8(61), 1987(63) & 1996-8(64) |
| Canada | Canada, <1967(65) |
| Chile | Chile 1967-8 (Santiago & rural)(61) & 1983 (Santiago) (66) |
| Colombia | AMR, excluding the Caribbean |
| Costa Rica | AMR, excluding the Caribbean |
| Cuba | Caribbean |
| Dominican Republic | Caribbean |
| Ecuador | AMR, excluding the Caribbean |
| El Salvador | AMR, excluding the Caribbean |
| French Guiana | Caribbean |
| Grenada | Caribbean |
| Guadeloupe | Caribbean |
| Guatemala | AMR, excluding the Caribbean |
| Guyana | Caribbean |
| Haiti | Haiti, 2003(70) |
| Honduras | AMR, excluding the Caribbean |
| Jamaica | Jamaica, 1967-8 (Kingston & rural)(61) |
| Martinique | Caribbean |
| Mexico | Mexico, 1987-88(67) & 1989(68) |

| | |
|---|---|
| Nicaragua | AMR, excluding the Caribbean |
| Panama | Panama 1967-8 (Panama City & rural)(61) |
| Paraguay | AMR, excluding the Caribbean |
| Peru | Peru, 1967-8 (Lima & rural)(61) & 2003(69) |
| Puerto Rico | USA (Atlanta and Houston), <1967(65) |
| Saint Lucia | Caribbean |
| Saint Vincent and the Grenadines | Caribbean |
| Suriname | Caribbean |
| Trinidad and Tobago | Trinidad 1966-7(71), 1967-8 (Port au Spain & rural)(61) |
| USA | USA <1967 (Atlanta & Houston)(65) |
| US Virgin Islands | USA <1967 (Atlanta & Houston)(65) |
| Uruguay | Uruguay, 1967-7 (urban and rural)(61) |
| Venezuela | AMR, excluding the Caribbean |
| **Eastern Mediterranean** | |
| Afghanistan | EMR |
| Bahrain | Bahrain, 1981(72) |
| Djibouti | EMR |
| Egypt | Egypt (Cairo), 1973(73) |
| Iran | Iran, 1993-95(74) |
| Iraq | EMR |
| Jordan | Jordan, 1982-3(75) |
| Kuwait | Kuwait, <1978(76) |
| Lebanon | Lebanon, 1980-81(77) |
| Libya | EMR |
| Morocco | Morocco, 1969-1970(78) |
| Sudan | Sudan (Khartoum), 2015-16(83) & 2016(84) |
| Oman | EMR |

| | |
|---|---|
| Pakistan | Pakistan, <1997(79), 1999-2004(80) |
| Qatar | EMR |
| Saudi Arabia | Saudi Arabia, 1989(81) & 1992-3(82) |
| Somalia | EMR |
| State of Palestine | EMR |
| Syrian Arab Republic | EMR |
| Tunisia | Tunisia, <1970(85) |
| United Arab Emirates | EMR |
| Yemen | Yemen, 1985(86) & 2002-3(87) |
| **Europe** | |
| Albania | EUR |
| Armenia | EUR |
| Austria | EUR |
| Azerbaijan | EUR |
| Belarus | EUR |
| Belgium | EUR |
| Bosnia and Herzegovina | EUR |
| Bulgaria | EUR |
| Channel Islands | EUR |
| Croatia | EUR |
| Cyprus | EUR |
| Czech Republic | Czech Republic, <1967(65) & 1984 (Prague) (88) |
| Denmark | Denmark, <1967(65) &1983(89) |
| Estonia | EUR |
| Finland | Finland, 1979(92) |
| France | France, <1967(65) |
| Georgia | EUR |

| | |
|---|---|
| Germany | East Germany, 1990(90) |
| Greece | EUR |
| Hungary | EUR |
| Iceland | EUR |
| Ireland | EUR |
| Israel | EUR |
| Italy | EUR |
| Kazakhstan | EUR |
| Kyrgyzstan | Kyrgyzstan, 1968-70(93) & 2001(94) |
| Latvia | EUR |
| Lithuania | EUR |
| Luxembourg | EUR |
| Malta | EUR |
| Montenegro | EUR |
| Netherlands | EUR |
| Norway | EUR |
| Poland | Poland, 1969(95), 1973(95), 1979 (urban)(96), 1979 (rural)(96), 1982 (urban)(95), 1982 (rural)(95) |
| Portugal | EUR |
| Moldova | EUR |
| Romania | Romania, <1989(97) |
| Russia | EUR |
| Serbia | EUR |
| Slovakia | EUR |
| Slovenia | EUR |
| Spain | Spain, 1969-71(98) |
| Sweden | EUR |
| Switzerland | Switzerland, 1985(99) |

| | |
|---|---|
| Macedonia | EUR |
| Tajikistan | EUR |
| Turkey | Turkey, 1998(100), 2003-04(101) & 2005(102) |
| Turkmenistan | EUR |
| Ukraine | EUR |
| United Kingdom | England, <1967(65) & 1986-87(91) |
| Uzbekistan | EUR |
| **South East Asia** | |
| Bangladesh | Bangladesh, 2004-5(103) |
| Bhutan | SEAR |
| India | India, 1968 (urban & rural Delhi)(104), 1972-3 (Chandrigarh & Lucknow)(104), 1976 (Calcutta)(105), <1987 (Delhi)(106), <1990 (Delhi)(107), 1999-2000 (urban and rural Vellore)(108), 2016 (Kerala) (109), 2017(110) |
| Indonesia | Indonesia, 2007 (*S Reef, personal communication)* |
| Korea, Democratic People's Republic | SEAR |
| Maldives | SEAR |
| Myanmar | SEAR |
| Nepal | Nepal, 2008(111) |
| Sri Lanka | SEAR |
| Thailand | Thailand, 1978(112) |
| Timor-Leste | SEAR |
| **Western Pacific** | |
| Australia | Australia, <1967(65) |
| Brunei Darussalam | WPR, excluding China & Australia |
| Cambodia | WPR, excluding China & Australia |
| China | China, 1979-80(123) |
| China (Hong Kong) | China, 1979-80(123) |

| | |
|---|---|
| China (Macao) | China, 1979-80(123) |
| Fiji | Fiji, <1973(113) |
| French Polynesia | WPR, excluding China & Australia |
| Guam | WPR, excluding China & Australia |
| Japan | Japan, <1967 (Sapporo &Ohtsu)(65) |
| Kiribati | WPR, excluding China & Australia |
| Laos | Laos, 2014 (114) |
| Malaysia | Malaysia, <1972(115) |
| Micronesia (Fed. States) | WPR, excluding China & Australia |
| Mongolia | WPR, excluding China & Australia |
| New Caledonia | WPR, excluding China & Australia |
| New Zealand | Australia, <1967(65) |
| Papua New Guinea | WPR, excluding China & Australia |
| Philippines | WPR, excluding China & Australia |
| Republic of Korea | WPR, excluding China & Australia |
| Samoa | WPR, excluding China & Australia |
| Singapore | Singapore, 1975-9(116) |
| Solomon Islands | WPR, excluding China & Australia |
| Taiwan | WPR, excluding China & Australia |
| Tonga | WPR, excluding China & Australia |
| Vanuatu | WPR, excluding China & Australia |
| Vietnam | Central Vietnam, 2009-2010(119) |

### D: Equations for the CRS incidence and annual numbers of CRS cases

For a given model run, *j*, out of the 1000 model runs, the number of CRS cases per 100,000 livebirths for a given country, *c*, for each year *y* during 1996-2019 was calculated using the following equation

$$I^{B}_{CRS,c,j}(A_{15-49}, y) = \frac{N_{CRS,c,j}(A_{15-49}, y)}{\sum_{a=15}^{49} f(a,y) N_w(a,y)} \times 100{,}000$$

Here, the denominator is the number of births in year *y* for all women aged 15-49 years, calculated using the fertility rate (*f(a,y)*) and population size ($N_w(a,y)$) of women aged *a* in year *y* in the UN population data and the numerator ($N_{CRS,c,j}(A_{15-49}, y)$) is the estimated number of CRS cases born to women aged 15- 49 years in year *y* for the $j^{th}$ set of parameter values. The latter number ($N_{CRS,c,j}(A_{15-49}, y)$) was calculated by summing the daily number of CRS cases born to women aged 15-49 years, as follows:

$$N_{CRS,c,j}(A_{15-49}, y) = \sum_{t=1}^{365} \sum_{a=15}^{49} \frac{r_{CRS,j} s_{w,j}(a,t) f(a,y) N_w(a,y)(1 - e^{-112\lambda_o(t)})}{365}$$

Here, $s_{w,j}(a,t)$ is the modelled proportion of women aged *a* on day *t* that are susceptible, $\lambda_{o,j}(t)$ is the daily model-generated force of infection among women on day t, and $r_{CRS,j}$ is the risk of a newborn of a mother infected during the first 16 weeks of pregnancy having CRS for the $j^{th}$ bootstrap set of values. As elsewhere(26, 124, 125), we assume that infection during the first 16 weeks of pregnancy carries an average 65% (95% range: (47,88%)) risk of the newborn having CRS.

*Regional and global estimates*

The regional median and 95% CI of the CRS incidence per 100,000 live births was calculated using country-specific estimates (see above), weighted by the population size.

The equation for the $j^{th}$ model run of the average CRS incidence per 100,000 live births for the $N$ countries in a given region was as follows:

$$\frac{\sum_{c=1}^{N} I_{CRS,c,j}^{B}(A_{15-49}, y) P_c(y)}{\sum_{c=1}^{N} P_c(y)}$$

where $I_{CRS,c,j}^{B}(A_{15-49}, y)$ is defined above and $P_c(y)$ is the population size of country $c$ in year $y$. As previously, given China's large population size, the regional incidence for WPR was calculated with and without excluding China, .

We summed the annual numbers for the $j^{th}$ set of bootstrap parameter values for each country in the region to obtain the corresponding regional totals, which were summed to obtain the global burden (i.e. including all countries). These calculations were repeated for each of the 1000 combinations of parameter values.

The 95% CI of the national, regional and global numbers of CRS cases were approximated by the 95% range of the corresponding 1000 values. The country-specific central value was taken as the median from 1000 model runs if the country had either no or >1 seroprevalence dataset. Otherwise, it was taken as that derived from the estimated prevaccination force of infection from the observed data.

#### E. Estimates of the pre-vaccination force of infection

Table E: Summary of the additional datasets that were identified since the previous systematic review, best-fitting values for the force of infection and (where appropriate) the sensitivity of the antibody assay for each catalytic model before the introduction of RCV. The values in parentheses reflect the 95% confidence intervals, obtained by bootstrapping. Several of the estimates were published in an interim update of the literature review(126) and are included here for completeness.

| **Country, year of study** | **Study population** | **Sample size (no. of age groups)** | **Lab test (cut-off)** | **Cata-lytic model** | **Force of infection (/1000/year)** | | **Sensitivity (%)** | **Loglike-lihood deviance (deg of freedom)** | **Selected model** |
|---|---|---|---|---|---|---|---|---|---|
| | | | | | **<13 yr olds** | **≥13 yr olds** | | | |
| **African region** | | | | | | | | | |
| Algeria, 2005-7(32) | Women of child-bearing age | 834(6) | ELISA, 10IU | A | 0 (0,942) | 503 (0,867) | 69 (66,100) | 2 (3) | |
| | | | | B | 93 (74,99) | 0 (0,12) | -- | 3 (4) | |
| | | | | C | 192 (93,933) | 192 (93,933) | 69 (66,74) | 3 (4) | |
| | | | | D | 36 (33,39) | 36 (33,39) | -- | 32 (5) | D |
| Burkina Faso, 2007-8(34) | Pregnant F | 341(4) | ELISA | A | 0 (0,915) | 828 (0,1000) | 96 (94,100) | 2(1) | B |
| | | | | B | 242 (135,282) | 3 (0,128) | - | 2(2) | |
| | | | | C | 235 (139,990) | 235 (139,990) | 96 (93,99) | 2(2) | |
| | | | | D | 126 (108,154) | 126 (108,154) | - | 10(3) | |
| Cameroon (Bafoussam), 2016(35) | pregnant women | 91(5) | ELISA, IgG Index >=1.00 | A | 212 (109,978) | 4 (0,836) | 100 (90,100) | 4 (2) | |
| | | | | B | 212 (109,300) | 4 (0,96) | -- | 4 (3) | |
| | | | | C | 999 (94,999) | 999 (94,999) | 93 (88,100) | 4 (3) | |
| | | | | D | 102 (78,151) | 102 (78,151) | -- | 6 (4) | A |

| Country, year of study | Study population | Sample size (no. of age groups) | Lab test (cut-off) | Cata-lytic model | Force of infection (/1000/year) | | Sensitivity (%) | Loglike-lihood deviance (deg of freedom) | Selected model |
|---|---|---|---|---|---|---|---|---|---|
| | | | | | <13 yr olds | ≥13 yr olds | | | |
| Cameroon (Yaounde), <2018(36) | Pregnant women at ANCs | 400(6) | ELISA ?, 10IU | A | 153 (33,593) | 51 (0,1000) | 100 (91,100) | 2 (3) | |
| | | | | B | 153 (95,215) | 51 (0,123) | -- | 2 (4) | |
| | | | | C | 149 (102,999) | 149 (102,999) | 95 (91,100) | 2 (4) | |
| | | | | D | 106 (93,123) | 106 (93,123) | -- | 4 (5) | B |
| Democratic Republic of the Congo (Kikwit), 2008-9(27) | Pregnant F | 254 (5) | ELISA, ≥10IU | A | 145 (103,632) | 27 (0,75) | 100 (89,100) | 5(2) | B |
| | | | | B | 145 (105,189) | 27 (0,69) | - | 5(3) | |
| | | | | C | 999 (83,999) | 999 (83,999) | 89 (85,100) | 6(3) | |
| | | | | D | 86 (74,103) | 86 (74,103) | - | 10(4) | |
| Democratic Republic of the Congo (Mikalayi), 2008-9(27) | Pregnant F | 206 (5) | ELISA, ≥10IU | A | 0 (0,466) | 557 (0,992) | 82 (77,100) | 0(2) | B |
| | | | | B | 103 (66,138) | 23 (0,68) | - | 1(3) | |
| | | | | C | 125 (67,969) | 125 (67,969) | 84 (76,99) | 1(3) | |
| | | | | D | 63 (54,77) | 63 (54,77) | - | 6(4) | |

| Country, year of study | Study population | Sample size (no. of age groups) | Lab test (cut-off) | Cata-lytic model | Force of infection (/1000/year) | | Sensitivity (%) | Loglike-lihood deviance (deg of freedom) | Selected model |
|---|---|---|---|---|---|---|---|---|---|
| | | | | | <13 yr olds | ≥13 yr olds | | | |
| Democratic Republic of the Congo (Tshikapa), 2008-9(27) | Pregnant F | 182 (5) | ELISA, ≥10IU | A | 128 (75,913) | 20 (0,248) | 100 (82,100) | 58 (0,187) | B |
| | | | | B | 128 (84,169) | 20 (0,73) | - | 58 (0,180) | |
| | | | | C | 168 (77,999) | 168 (77,999) | 86 (80,100) | 58 (0,200) | |
| | | | | D | 76 (62,94) | 76 (62,94) | - | 202 (163,232) | |
| Democratic Republic of the Congo (Vange), 2008-9(27) | Pregnant F | 255 (5) | ELISA, ≥10IU | A | 132 (90,252) | 32 (0,75) | 100 (91,100) | 75 (0,165) | B |
| | | | | B | 132 (91,178) | 32 (0,72) | - | 75 (0,164) | |
| | | | | C | 968 (77,999) | 968 (77,999) | 87 (84,100) | 0 (0,200) | |
| | | | | D | 83 (71,97) | 83 (71,97) | - | 187 (157,211) | |
| Ethiopia (Amhara), 2015-17(120) | Pregnant women | 600(5) | EIA, >10IU | A | 113 (0,425) | 19 (10,815) | 100 (78,100) | 0 (2) | |
| | | | | B | 108 (72,136) | 17 (0,51) | -- | 2 (3) | |
| | | | | C | 108 (76,837) | 108 (76,837) | 88 (78,93) | 1 (3) | |
| | | | | D | 60 (54,67) | 60 (54,67) | -- | 9 (4) | B |

| Country, year of study | Study population | Sample size (no. of age groups) | Lab test (cut-off) | Cata -lytic mod el | Force of infection (/1000/year) | | Sensitivity (%) | Loglike-lihood deviance (deg of freedom) | Selected model |
|---|---|---|---|---|---|---|---|---|---|
| | | | | | <13 yr olds | ≥13 yr olds | | | |
| Ethiopia (Hawassa), 2016(42) | Pregnant women at ANCs | 422(5) | ELISA | A | 0 (0,987) | 985 (0,1000) | 86 (83,97) | 2 (2) | |
| | | | | B | 159 (130,179) | 0 (0,26) | -- | 3 (3) | |
| | | | | C | 606 (142,989) | 606 (142,989) | 84 (83,91) | 3 (3) | |
| | | | | D | 73 (70,89) | 73 (70,89) | -- | 18 (4) | D |
| Kenya (Eldoret), 2005(121) | Pregnant F | 437(4) | EIA, ≥10IU | A | 140 (0,219) | 147 (24,875) | 97 (93,100) | 0(1) | B |
| | | | | B | 154 (91,223) | 66 (1,152) | - | 1(2) | |
| | | | | C | 142 (107,950) | 142 (107,950) | 97 (93,100) | 0(2) | |
| | | | | D | 113 (100,131) | 113 (100,131) | - | 2(3) | |
| Namibia, 2010(49) | Pregnant women attending ANCS | 2040(6) | EIA, OD>0.2 | A | 146 (0,167) | 24 (10,705) | 100 (90,100) | 7 (3) | B |
| | | | | B | 146 (127,167) | 24 (10,39) | -- | 7 (4) | |
| | | | | C | 168 (126,894) | 168 (126,894) | 91 (89,93) | 11 (4) | |
| | | | | D | 81 (76,86) | 81 (76,86) | -- | 60 (5) | |

| Country, year of study | Study population | Sample size (no. of age groups) | Lab test (cut-off) | Cata-lytic model | Force of infection (/1000/year) | | Sensitivity (%) | Loglike-lihood deviance (deg of freedom) | Selected model |
|---|---|---|---|---|---|---|---|---|---|
| | | | | | <13 yr olds | ≥13 yr olds | | | |
| Nigeria (Kaduna), 2011-12(53) | pregnant women attending ANCs | 400(6) | ELISA | A | 681 (0,985) | 0 (0,1000) | 97 (95,100) | 1 (3) | |
| | | | | B | 268 (201,313) | 0 (0,74) | -- | 1 (4) | |
| | | | | C | 962 (185,999) | 962 (185,999) | 97 (95,99) | 1 (4) | |
| | | | | D | 137 (118,168) | 137 (118,168) | -- | 13 (5) | D |
| Nigeria (Ilorin), 2012(54) | women of child-bearing age attending general outpatients department | 285(4) | ELISA | A | 160 (0,544) | 39 (0,663) | 100 (93,100) | 1 (1) | |
| | | | | B | 160 (104,226) | 39 (0,104) | -- | 1 (2) | |
| | | | | C | 158 (96,995) | 158 (96,995) | 94 (90,100) | 2 (2) | |
| | | | | D | 99 (85,121) | 99 (85,121) | -- | 5 (3) | B |
| Nigeria (Maiduguri), 2013(55) | Pregnant ANC attendees | 90(5) | ELISA | A | 118 (0,978) | 717 (0,999) | 84 (77,99) | 5 (2) | D |
| | | | | B | 143 (78,184) | 0 (0,65) | -- | 5 (3) | |
| | | | | C | 311 (91,999) | 311 (91,999) | 83 (77,95) | 5 (3) | |
| | | | | D | 73 (57,96) | 73 (57,96) | -- | 11 (4) | |

| Country, year of study | Study population | Sample size (no. of age groups) | Lab test (cut-off) | Cata-lytic model | Force of infection (/1000/year) | | Sensitivity (%) | Loglike-lihood deviance (deg of freedom) | Selected model |
|---|---|---|---|---|---|---|---|---|---|
| | | | | | <13 yr olds | ≥13 yr olds | | | |
| Nigeria (Kaduna), 2015(56) | Pregnant women attending ANC | 900(8) | ELISA | A | 132 (14,986) | 42 (2,1000) | 72 (62,100) | 7 (5) | |
| | | | | B | 72 (55,85) | 9 (0,24) | -- | 7 (6) | |
| | | | | C | 135 (71,998) | 135 (71,998) | 66 (61,77) | 7 (6) | |
| | | | | D | 40 (36,44) | 40 (36,44) | -- | 23 (7) | B |
| Nigeria (Keffi), <2016 (127) | Pregnant women at ANC | 220(5) | ELISA, IgG index 1.0 | A | 359 (0,930) | 0 (0,629) | 12 (10,100) | 2 (2) | |
| | | | | B | 10 (1,14) | 0 (0,8) | -- | 2 (3) | |
| | | | | C | 891 (4,995) | 891 (4,995) | 12 (8,100) | 2 (3) | |
| | | | | D | 5 (3,7) | 5 (3,7) | -- | 4 (4) | D – drop |
| South Africa (Soweto), 2014-16(59) | Pregnant women during labour or within 24 hours after delivery | 552(3) | ELISA, ≥11IU | A | 249 (141,382) | 71 (0,202) | 100 (100,100) | 4 (0) | B |
| | | | | B | 249 (141,364) | 71 (0,195) | -- | 4 (1) | |
| | | | | C | 212 (143,963) | 212 (143,963) | 99 (98,100) | 5 (1) | |
| | | | | D | 160 (139,199) | 160 (139,199) | -- | 6 (2) | |

| Country, year of study | Study population | Sample size (no. of age groups) | Lab test (cut-off) | Cata-lytic model | Force of infection (/1000/year) | | Sensitivity (%) | Loglike-lihood deviance (deg of freedom) | Selected model |
|---|---|---|---|---|---|---|---|---|---|
| | | | | | <13 yr olds | ≥13 yr olds | | | |
| Tanzania (Mwanza), 2012-13(122) | Pregnant F | 342 (3) | EIA, ≥10IU | A | 0 (0,226) | 544 (20,760) | 96 (93,100) | 0(0) | B |
| | | | | B | 142 (74,216) | 79 (6,190) | - | 1(1) | |
| | | | | C | 135 (105,265) | 135 (105,265) | 98 (93,100) | 0(1) | |
| | | | | D | 115 (99,137) | 115 (99,137) | - | 1(2) | |
| **American region** | | | | | | | | | |
| Chile (Santiago), 1983(66) | Pregnant women | 812(5) | ELISA | A | 0 (0,283) | 866 (8,799) | 95 (94,100) | 2 (2) | B |
| | | | | B | 182 (131,236) | 45 (0,104) | -- | 2 (3) | |
| | | | | C | 164 (121,971) | 164 (121,971) | 96 (94,100) | 2 (3) | |
| | | | | D | 115 (105,129) | 115 (105,129) | -- | 9 (4) | |
| **Eastern Mediterranean region** | | | | | | | | | |
| Egypt (Cairo), 1973(73) | Healthy population (well-baby clinic attendees, schoolchildren, students & women at gynaecology & obstetrics | 402(7) | HAI | A | 1000 (631,1000) | 121 (71,171) | 88 (85,91) | 27 (4) | B |
| | | | | B | 175 (150,210) | 49 (14,86) | -- | 64 (5) | |
| | | | | C | 1000 (637,1000) | 1000 (637,1000) | 88 (85,91) | 27 (5) | |
| | | | | D | 148 (129,175) | 148 (129,175) | -- | 70 (6) | |

| Country, year of study | Study population | Sample size (no. of age groups) | Lab test (cut-off) | Cata-lytic model | Force of infection (/1000/year) | | Sensitivity (%) | Loglike-lihood deviance (deg of freedom) | Selected model |
|---|---|---|---|---|---|---|---|---|---|
| | | | | | <13 yr olds | ≥13 yr olds | | | |
| Sudan (Khartoum), 2015-16(83) | General population | 447(6) | ELISA | A | 123 (100,147) | 1000 (714,1000) | 95 (92,98) | 10 (3) | B |
| | | | | B | 129 (108,150) | 51 (24,99) | -- | 17 (4) | |
| | | | | C | 136 (113,159) | 136 (113,159) | 96 (92,99) | 16 (4) | |
| | | | | D | 107 (92,126) | 107 (92,126) | -- | 27 (5) | |
| Sudan (Khartoum), 2016(84) | Healthy pregnant women | 92(3) | EIA, >10IU | A | 194 (0,992) | 8 (0,914) | 100 (88,100) | 0 (0) | B |
| | | | | B | 195 (53,274) | 8 (0,136) | -- | 0 (1) | |
| | | | | C | 270 (82,945) | 270 (82,945) | 92 (88,100) | 0 (1) | |
| | | | | D | 86 (67,125) | 86 (67,125) | -- | 3 (2) | |
| **Europe** | | | | | | | | | |
| Czech (Prague), 1984(88) | Pregnant women | 850(4) | HI test | A | 130 (42,181) | 57 (18,241) | 100 (94,100) | 1 (1) | A |
| | | | | B | 130 (80,177) | 57 (16,110) | -- | 1 (2) | |
| | | | | C | 111 (89,750) | 111 (89,750) | 97 (91,100) | 2 (2) | |
| | | | | D | 92 (85,101) | 92 (85,101) | -- | 4 (3) | |

| Country, year of study | Study population | Sample size (no. of age groups) | Lab test (cut-off) | Cata-lytic model | Force of infection (/1000/year) | | Sensitivity (%) | Loglike-lihood deviance (deg of freedom) | Selected model |
|---|---|---|---|---|---|---|---|---|---|
| | | | | | <13 yr olds | ≥13 yr olds | | | |
| Kyrgyzstan, 1968-70(93) | Donor sites and maternity hospitals | 1629(3) | HI test | A | 148 (28,176) | 70 (17,240) | 95 (91,100) | 0 (0) | A |
| | | | | B | 140 (114,169) | 30 (11,51) | -- | 0 (1) | |
| | | | | C | 125 (101,180) | 125 (101,180) | 93 (90,96) | 0 (1) | |
| | | | | D | 82 (77,88) | 82 (77,88) | -- | 22 (2) | |
| Poland, 1969(95) | girls and women | 1087(6) | ? | A | 182 (153,217) | 320 (136,818) | 96 (95,99) | 4 (3) | B |
| | | | | B | 174 (151,199) | 85 (43,139) | -- | 9 (4) | |
| | | | | C | 186 (158,221) | 186 (158,221) | 97 (95,99) | 5 (4) | |
| | | | | D | 151 (138,165) | 151 (138,165) | -- | 18 (5) | |
| Poland, 1973(95) | girls and women | 1066(5) | ? | A | 161 (137,189) | 620 (289,1000) | 95 (93,97) | 3 (2) | B |
| | | | | B | 157 (138,177) | 91 (49,145) | -- | 12 (3) | |
| | | | | C | 170 (147,197) | 170 (147,197) | 97 (94,99) | 9 (3) | |
| | | | | D | 141 (130,154) | 141 (130,154) | -- | 17 (4) | |
| Poland (urban), 1979(96) | Randomly selected sites | 866(10) | HI test, 1:10 | A | 149 (128,175) | 135 (92,231) | 99 (97,100) | 13 (7) | A |
| | | | | B | 148 (129,171) | 101 (65,153) | -- | 14 (8) | |
| | | | | C | 148 (130,175) | 148 (130,175) | 99 (97,100) | 13 (8) | |
| | | | | D | 137 (122,153) | 137 (122,153) | -- | 16 (9) | |

| Country, year of study | Study population | Sample size (no. of age groups) | Lab test (cut-off) | Cata -lytic mod el | Force of infection (/1000/year) | | Sensitivity (%) | Loglike-lihood deviance (deg of freedom) | Selected model |
|---|---|---|---|---|---|---|---|---|---|
| | | | | | <13 yr olds | ≥13 yr olds | | | |
| Poland (rural), 1979(96) | Randomly selected sites | 763(10) | HI test, 1:10 | A | 126 (104,145) | 314 (103,1000) | 97 (95,100) | 14 (7) | B |
| | | | | B | 128 (110,148) | 117 (76,186) | -- | 17 (8) | |
| | | | | C | 132 (117,152) | 132 (117,152) | 99 (97,100) | 16 (8) | |
| | | | | D | 126 (113,141) | 126 (113,141) | -- | 17 (9) | |
| Poland (urban), 1982(95) | Healthy population | 666(8) | HI test | A | 172 (150,211) | 95 (58,1000) | 100 (96,100) | 6 (5) | B |
| | | | | B | 172 (150,197) | 95 (50,169) | -- | 6 (6) | |
| | | | | C | 179 (151,224) | 179 (151,224) | 98 (95,100) | 7 (6) | |
| | | | | D | 158 (142,178) | 158 (142,178) | -- | 9 (7) | |
| Poland (rural), 1982(95) | Healthy population | 545(8) | HI test | A | 157 (135,187) | 106 (64,1000) | 100 (95,100) | 15 (5) | B |
| | | | | B | 157 (135,183) | 106 (60,192) | -- | 15 (6) | |
| | | | | C | 156 (134,199) | 156 (134,199) | 99 (95,100) | 16 (6) | |
| | | | | D | 148 (132,168) | 148 (132,168) | -- | 16 (7) | |

| Country, year of study | Study population | Sample size (no. of age groups) | Lab test (cut-off) | Cata-lytic model | Force of infection (/1000/year) | | Sensitivity (%) | Loglike-lihood deviance (deg of freedom) | Selected model |
|---|---|---|---|---|---|---|---|---|---|
| | | | | | <13 yr olds | ≥13 yr olds | | | |
| Spain, 1969-71(98) | Healthy population at large | 1076(9) | HI test | A | 378 (300,487) | 284 (0,1000) | 89 (86,92) | 15 (6) | C |
| | | | | B | 204 (188,223) | 0 (0,4) | -- | 41 (7) | |
| | | | | C | 378 (303,489) | 378 (303,489) | 89 (86,92) | 15 (7) | |
| | | | | D | 158 (143,179) | 158 (143,179) | -- | 163 (8) | |
| Switzerland, 1985(99) | Sera submitted for diagnostic testing unrelated to rubella | 736(28) | ELISA, 10IU | A | 51 (37,67) | 224 (153,318) | 92 (89,95) | 50 (25) | B |
| | | | | B | 64 (50,80) | 64 (46,88) | -- | 75 (26) | |
| | | | | C | 78 (66,92) | 78 (66,92) | 95 (92,99) | 68 (26) | |
| | | | | D | 64 (58,72) | 64 (58,72) | -- | 75 (27) | |
| **South East Asia** | | | | | | | | | |
| India (Kerala), 2016(109), | Pregnant women | 70(3) | ELISA, 15IU | A | 0 (0,993) | 838 (0,972) | 95 (90,100) | 1 (0) | D |
| | | | | B | 229 (0,340) | 0 (0,448) | -- | 1 (1) | |
| | | | | C | 343 (109,996) | 343 (109,996) | 94 (90,100) | 1 (1) | |
| | | | | D | 120 (90,196) | 120 (90,196) | -- | 2 (2) | |

| Country, year of study | Study population | Sample size (no. of age groups) | Lab test (cut-off) | Cata-lytic model | Force of infection (/1000/year) | | Sensitivity (%) | Loglike-lihood deviance (deg of freedom) | Selected model |
|---|---|---|---|---|---|---|---|---|---|
| | | | | | <13 yr olds | ≥13 yr olds | | | |
| India, 2017(110) | Pregnant women attending ANC clinic or womens hospital | 1800(5) | ELISA, >10iu | A | 344 (0,994) | 0 (0,966) | 86 (83,100) | 1 (2) | D |
| | | | | B | 151 (135,159) | 0 (0,14) | -- | 1 (3) | |
| | | | | C | 854 (179,994) | 854 (179,994) | 85 (83,87) | 1 (3) | |
| | | | | D | 73 (69,78) | 73 (69,78) | -- | 45 (4) | |
| Indonesia, 2007 (*S Reef, personal communication, March 2015*) | General population | 11320 (10) | ? | A | 135 (119,148) | 62 (32,97) | 93 (91,97) | 3(7) | B |
| | | | | B | 127 (120,135) | 22 (18,26) | - | 12(8) | |
| | | | | C | 115 (106,126) | 115 (106,126) | 91 (90,92) | 12(8) | |
| | | | | D | 61 (60,63) | 61 (60,63) | - | 462(9) | |
| **Western Pacific** | | | | | | | | | |
| Cambodia, 2012(128) | Nationwide cross-section of women aged 15-39 years | 2154(5) | ElISA (OD >0.2 | A | 64 (0,100) | 191 (71,421) | 80 (77,87) | 1(2) | B |
| | | | | B | 76 (65,89) | 33 (21,44) | -- | 8(3) | |
| | | | | C | 91 (74,116) | 91 (74,116) | 84 (79,89) | 3(3) | |
| | | | | D | 54 (51,58) | 54 (51,58) | -- | 23(4) | |

| Country, year of study | Study population | Sample size (no. of age groups) | Lab test (cut-off) | Cata-lytic model | Force of infection (/1000/year) | | Sensitivity (%) | Loglike-lihood deviance (deg of freedom) | Selected model |
|---|---|---|---|---|---|---|---|---|---|
| | | | | | <13 yr olds | ≥13 yr olds | | | |
| Laos, 2014 (114)* | General population aged >21 years(i.e. excluding the age range vaccinated in the SIA in 2011) | 1013 (45) | ELISA, >10IU | A | 90 (38,141) | 18 (9,127) | 100 (82,100) | 60 (42) | B |
| | | | | B | 90 (69,111) | 18 (8,29) | - | 60 (43) | |
| | | | | C | 80 (58,129) | 80 (58,129) | 85 (79,91) | 61 (43) | |
| | | | | D | 44 (40,47) | 44 (40,47) | - | 78 (44) | |

* The methods for calculating the force of infection for Laas are identical to those presented in (114), except that, for consistency with the estimates for the other countries, the force of infection is assumed to differ between those aged ≤13 years and those aged >13 years.

#### F. Estimates of the CRS incidence

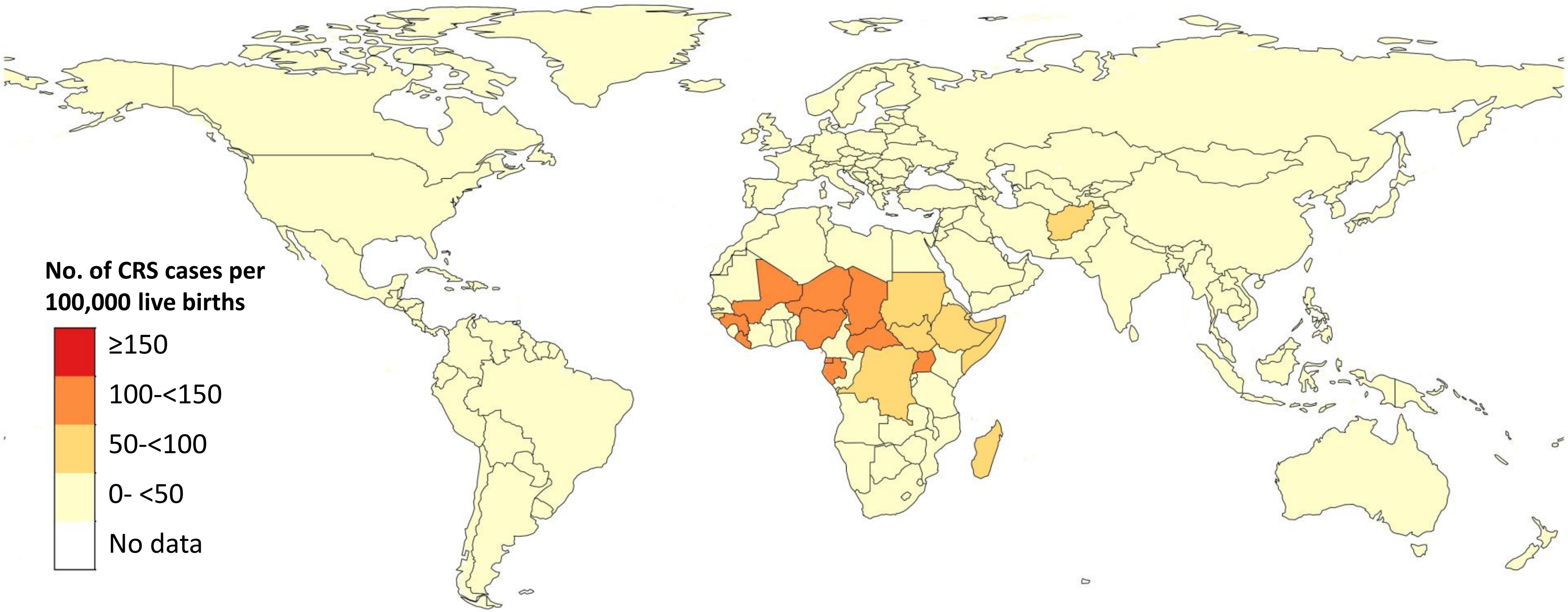


Figure S1: Average estimates of the number of CRS cases per 100,000 live births in 2019 for all countries.

Table F: The median CRS incidence per 100,000 live births and number of CRS cases born in each WHO region and worldwide in 1996, 2000, 2010 and 2019 and the percentage of the regional live births occurring in countries which had introduced RCV by these years. The numbers in parentheses reflect 95% confidence limits.

| | Year | % of the regional live births in countries with RCV | CRS incidence per 100,000 live births | Total number of CRS cases |
|---|---|---|---|---|
| **African region** | 1996 | 0.15 | 121 (64,211) | 29468 (14763,52921) |
| | 2000 | 0.14 | 121 (63,212) | 32073 (15935,57609) |
| | 2010 | 0.16 | 119 (62,211) | 38873 (19574,70294) |
| | 2019 | 41 | 64 (24,123) | 25454 (9193,48881) |
| **Eastern Mediterranean region** | 1996 | 12 | 62 (33,110) | 8440 (3976,15981) |
| | 2000 | 30 | 54 (24,109) | 8011 (2908,17387) |
| | 2010 | 45 | 28 (5,68) | 5514 (1116,13043) |
| | 2019 | 49 | 27 (4,67) | 5660 (873,14073) |
| **European region** | 1996 | 52 | 71 (21,176) | 8534 (2920,20403) |
| | 2000 | 68 | 40 (15,101) | 5682 (2092,13184) |
| | 2010 | 100 | 4 (1,18) | 319 (52,1545) |
| | 2019 | 100 | 1 (0,12) | 100 (0,957) |
| **Region of the Americas** | 1996 | 61 | 58 (26,112) | 11626 (5241,22202) |
| | 2000 | 90 | 12 (6,26) | 2657 (1170,5562) |
| | 2010 | 100 | <1 (0,3) | 1 (0,357) |
| | 2019 | 100 | <1 (0,1) | <1 (0,90) |
| **South East Asian region** | 1996 | 3 | 126 (30,247) | 50035 (10866,99711) |
| | 2000 | 3 | 123 (28,250) | 49404 (10379,101215) |
| | 2010 | 3 | 121 (26,245) | 45444 (9316,92325) |
| | 2019 | 100 | <1 (<1,8) | 51 (<1,1662) |
| **Western Pacific Region (excluding China)** | 1996 | 41 | 123 (59,234) | 11741 (5501,22734) |
| | 2000 | 40 | 113 (56,214) | 10925 (5235,19887) |
| | 2010 | 69 | 101 (50,199) | 10006 (4700,19258) |
| | 2019 | 100 | <1 (<1,44) | <1 (<1,3756) |
| **Western Pacific Region (including China)** | 1996 | 12 | 33 (16,61) | 12179 (5887,22940) |
| | 2000 | 12 | 31 (16,58) | 11486 (5569,20502) |
| | 2010 | 91 | 27 (13,52) | 10023 (4704,19261) |
| | 2019 | 100 | <1 (0,12) | <1 (0,3768) |
| **Global** | 1996 | 16 | - | 120687 (70405,191204) |
| | 2000 | 23 | - | 109353 (61616,179222) |
| | 2010 | 42 | - | 99665 (53554,165778) |
| | 2019 | 77 | - | 32460 (12794,59818) |

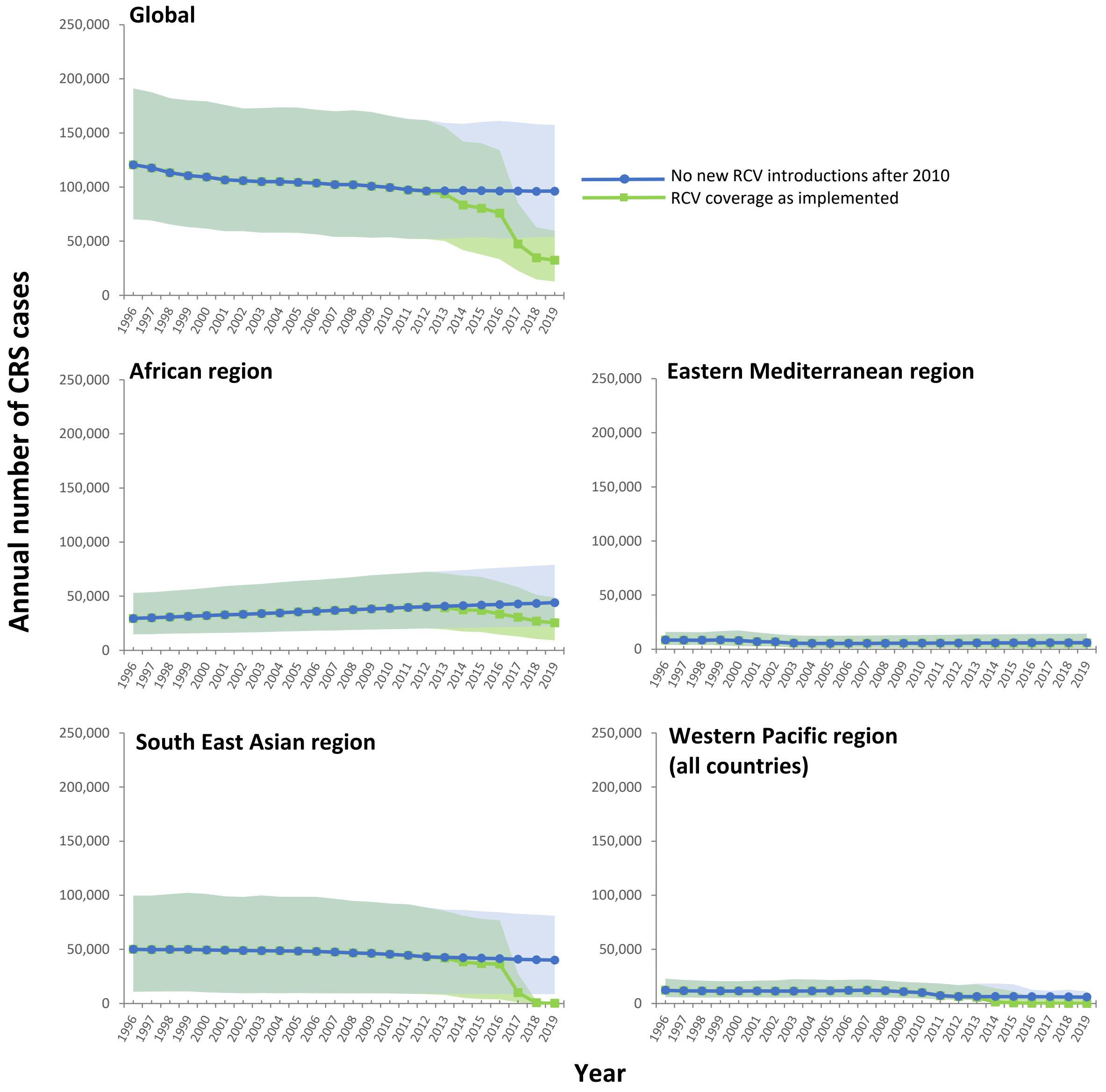


Figure S2: Estimates of the annual number of CRS cases during 1996-2019 globablly, in the African, Eastern Mediterranean, South East Asian regions, as calculated using the RCV coverage as implemented and the number that might have been seen if there had been no new introductions of RCV after 2010. The shaded areas show the 95% confidence intervals.
